# Baseline functional connectivity in resting state networks associated with depression and remission status after 16 weeks of pharmacotherapy: A CAN-BIND Report

**DOI:** 10.1101/2021.01.27.21250490

**Authors:** Gwen van der Wijk, Jacqueline K. Harris, Stefanie Hassel, Andrew D. Davis, Mojdeh Zamyadi, Stephen R. Arnott, Roumen Milev, Raymond W. Lam, Benicio N. Frey, Geoffrey B. Hall, Daniel J. Müller, Susan Rotzinger, Sidney H. Kennedy, Stephen C. Strother, Glenda M. MacQueen, Andrea B. Protzner

## Abstract

Understanding the neural underpinnings of major depressive disorder (MDD) and its treatment could improve treatment outcomes. While numerous studies have been conducted, findings are variable and large sample replications scarce. We aimed to replicate and extend altered functional connectivity findings in the default mode, salience and cognitive control networks (DMN, SN, and CCN respectively) associated with MDD and pharmacotherapy outcomes in a large, multi-site sample. Resting-state fMRI data were collected from 129 patients and 99 controls through the Canadian Biomarker Integration Network in Depression (CAN-BIND) initiative. Symptoms were assessed with the Montgomery-Åsberg Depression Rating Scale (MADRS). Connectivity was measured as correlations between four seeds (anterior and posterior DMN, SN and CCN) and all other brain voxels across participants. Partial least squares, a multivariate statistical technique, was used to compare connectivity prior to treatment between patients and controls, and between patients reaching remission early (MADRS ≤ 10 within 8 weeks), late (MADRS ≤ 10 within 16 weeks) or not at all. We replicated previous findings of altered connectivity in the DMN, SN and CCN in patients. In addition, baseline connectivity of the anterior/posterior DMN and SN seeds differentiated patients with different treatment outcomes. Weaker connectivity within the anterior DMN and between the anterior DMN and the SN and CCN characterised early remission; stronger connectivity within the SN and weaker connectivity between the SN and the DMN and CCN was related to late remission, of which the weaker SN – anterior DMN connectivity might specifically be associated with remission to dual pharmacotherapy; and connectivity strength between the posterior DMN and cingulate areas distinguished all three groups, with early remitters showing the strongest connections and non-remitters the weakest. The stability of these baseline patient differences was established in the largest single-site subsample of the data. Our replication and extension of altered connectivity within and between the DMN, SN and CCN highlighted previously reported and new differences between patients with MDD and controls, and revealed features that might predict remission prior to pharmacotherapy.

**Trial registration:** ClinicalTrials.gov: NCT01655706.

## Introduction

Major depressive disorder (MDD) is a prevalent and debilitating disorder. Although several effective treatments are available, including psychotherapy, pharmacotherapy and neurostimulation therapy, treatment outcomes vary greatly among patients [1, 2]. Many neuroimaging studies have investigated brain changes related to MDD pathology and antidepressant treatment, but while the results are promising, inconsistent findings have led to growing concerns about reproducibility [3]. To improve treatment outcomes for patients with MDD, a robust understanding of the characteristics associated with MDD and antidepressant treatment outcomes is essential.

Functional connectivity during rest, as captured by functional magnetic resonance imaging (fMRI) of the brain, has emerged as a potentially informative measure of the neural underpinnings of both the depressive state and its (successful) treatment [4–7]. Based on the more robust findings highlighted in meta-analyses and literature reviews, the three networks that have most consistently been linked to MDD pathology include the default mode network (DMN), the salience network (SN) and the cognitive control network (CCN) [4, 8]. Core regions of the DMN are the posterior cingulate cortex (PCC), precuneus, the bilateral angular gyrus and the medial prefrontal cortex (mPFC; [4]). In MDD, the anterior cingulate cortex (ACC) has also been found to be part of the DMN [9]. This network is generally thought to be related to internal processing in conscious participants ([4], see [10] for a discussion on DMN activity during unconscious states). The core regions of the SN include the bilateral insulae, the amygdalae, the temporal poles and the dorsal anterior cingulate cortex (dACC; [4]). It is involved in emotion processing and bottom-up attentional processes (e.g. monitoring for salient stimuli; [5]). Core CCN regions include the bilateral dorsolateral prefrontal cortex (dlPFC), frontal eye fields (FEF), dorsomedial prefrontal cortex (dmPFC) and the posterior parietal cortex (PPC), which together exert top-down control over cognitive and emotional processes [4]. Disturbed connectivity within and between these networks is thought to lead to the abnormal emotion processing and mood regulation in MDD [5, 11]. Given the frequency with which these networks are highlighted in previous work and their functional relevance to MDD pathology, we focused on these three resting state networks in our study.

A meta-analysis and literature review summarizing resting state fMRI connectivity alterations in patients with MDD compared to controls reported that MDD was characterized by stronger connectivity within the DMN, especially anteriorly, between the CCN and DMN, and between the anterior DMN and SN [4, 5]. In addition, weaker connectivity within the CCN, between the SN and the posterior DMN, and between the posterior DMN and CNN was found in patients with MDD compared to controls across studies [4, 5]. However, the studies included in these papers generally had small samples (typically < 30 patients) collected at a single site, and variable methodologies, limiting the generalizability of these findings. In addition, MDD itself is a heterogeneous disorder, which likely contributes to the variable findings [12, 13]. Several authors have called for large, multi-site datasets to increase replicability (e.g. [14, 15]).

Several studies have also investigated connectivity differences between patients with different treatment outcomes. Here we focus on findings related to pharmacotherapy, which is the most common first-line treatment for MDD. Three reviews indicate that findings often include the same three networks as mentioned above, most commonly showing a decrease in connectivity within the DMN, an increase in connectivity within the CCN and between frontal (CCN) areas and limbic (SN) regions with treatment [6–8]. However, the authors of these reviews emphasize that results vary greatly among studies, and that similar issues to those raised in the previous paragraph prevent robust conclusions and limit translational relevance [6–8]. Indeed, a recent meta-analysis was not able to detect any resting state connectivity patterns predictive of treatment response to pharmacotherapy [16]. They included only studies examining the association between symptom improvement with treatment and baseline brain-wide resting state connectivity at the voxel level, leaving six studies for their analysis. In addition to underlining the variability in findings to date, this highlights the scarcity of studies looking specifically at baseline fMRI connectivity predictors of treatment outcomes, which could be especially informative for clinical practice.

In the current study, we made use of a relatively large, multi-site resting state fMRI dataset to replicate and extend previous findings of connectivity alterations in patients with MDD compared to controls, and differences in connectivity prior to treatment among patients with differing outcomes. The dataset was collected as part of the Canadian Biomarker Integration Network in Depression 1 (CAN-BIND-1) study and included controls and patients diagnosed with MDD, who were treated for 16 weeks with escitalopram, and an add-on of aripiprazole after 8 weeks if symptoms decreased less than 50% [17, 18]. Depression symptoms were monitored using the Montgomery-Åsberg Depression Rating Scale (MADRS; [19]). Patients were divided into three groups: *early remitters* reached a score of ≤ 10 on the MADRS by week 8, *late remitters* reached this threshold after 16 weeks, and *non-remitters* did not reach remission by the end of the 16-week trial. Resting state fMRI data were recorded before the start of treatment. Connectivity between key nodes from the DMN, SN and CCN, and the rest of the brain was compared between groups using partial least squares (PLS; [20]). We expected to replicate previous patient-control differences in connectivity within and between the DMN, SN and CCN, and to find differences among patients with different treatment outcomes in these same networks.

## Methods

### Participants & treatment

Study participants included 108 controls and 200 patients (70 & 126 females, respectively) with a primary diagnosis of major depressive disorder (MDD), who were recruited from six academic health centres across Canada (University of British Columbia, University of Calgary, McMaster University, Centre for Addiction and Mental Health, Toronto General Hospital, Queen’s University) as part of the CAN-BIND-1 study [17, 18]. Briefly, patients were between 18-60 years of age, spoke sufficient English to complete the study, met the criteria for a major depressive episode according to the DSM-IV-TR, as assessed with the Mini International Neuropsychiatric Interview (MINI, [21]), had symptom scores of ≥ 24 on the Montgomery-Åsberg Depression Rating Scale ([MADRS]; [19]) and their current episode lasted 3 months or longer. Patients were excluded if they met the diagnostic criteria of bipolar-I or -II disorder or any other primary psychiatric or personality disorder (except for generalized anxiety disorder and social anxiety disorder), experienced psychotic symptoms in the current episode, had a history of neurological disorders, head trauma or other unstable medical conditions, had a high risk of suicide or a hypomanic switch, experienced substance dependence/abuse in the last 6 months, were currently pregnant or breastfeeding, showed previous non-response to four adequate pharmacotherapy interventions, had a previous unfavorable response to escitalopram or aripiprazole or had any contraindications to MRI. Patients who had been taking antidepressant medication prior to participation went through a washout period (lasting at least 5 half-lives). Controls were between 18-60 years of age, had no psychiatric or unstable physical health diagnosis, no history of neurological disorders, head trauma or other unstable medical conditions, and spoke sufficient English to complete the study.

Ethics approval was obtained from all participating centres. The ethics committees include: University of British Columbia Clinical Research Ethics Board (Vancouver); University of Calgary Conjoint Health Research Ethics Board (Calgary); University Health Network Research Ethics Board (Toronto); Centre for Addiction and Mental Health Research Ethics Board (Toronto); Hamilton Integrated Research Ethics Board (Hamilton); Queen’s University Health Sciences and Affiliated Teaching Hospitals Research Ethics Board (Kingston). The participants provided written, informed consent for all study procedures.

All patients were given escitalopram for the first 8 weeks of the study. The initial dose was 10mg/d, which was increased to 20mg/d if MADRS symptom scores did not drop by 20% after two weeks or 50% after four weeks. Patients who improved less than 50% over 8 weeks were given a flexibly dosed add-on of aripiprazole (2-10mg/d) for an additional 8 weeks. Patients who improved 50% or more remained on their effective dose of escitalopram. Treating psychiatrists could prescribe lower doses for patients who did not tolerate higher doses, and the use of non-psychotropic medications for stable conditions, non-prescription analgesics, supplementals, vitamins and oral contraceptives were allowed at the discretion of the study psychiatrist.

Depression symptoms were assessed with the MADRS, using the structured interview guide (SIGMA) to enhance reliability [22], every two weeks during treatment. For the group analyses, patients were assigned to one of three groups based on whether, and when, they reached remission, which was defined as having a MADRS score ≤ 10, roughly equivalent to the clinical consensus of a cut-off score of ≤ 7 on the Hamilton Rating Scale for Depression [23]. Remission status was chosen as the main outcome measure for this analysis because this is the ultimate treatment goal, as residual symptoms can lead to significant morbidity [24–26]. Patients who achieved remission at 8 weeks of treatment and maintained this at 16 weeks were considered early remitters (ER), patients who reached this threshold at 16 weeks were considered late remitters (LR), and patients whose MADRS scores were above 10 throughout the 16 weeks were considered non-remitters (NR). Patients who reached remission at 8 weeks, but then relapsed by week 16 (N = 6) were not included. An additional 64 participants were excluded due to; treatment not being initiated (N = 10), missing clinical data (N = 23) and missing fMRI data or poor fMRI data quality (N = 31), leaving a sample of 129 patients for analysis. From the control sample, 9 participants were excluded due to poor fMRI data quality. The characteristics of the patient and control groups, as well as ER, LR, and NR patient groups are presented in **Table 1** and **Table 2**, respectively. Statistical differences were assessed in Excel, and any missing individual data points were replaced by the average of their allocated group (e.g. controls or early remitters). The characteristics for which there were missing data points are marked in **Table 1** and **Table 2**, and for each of these, there were no more than three individuals with missing data per group.

**Table 1.**
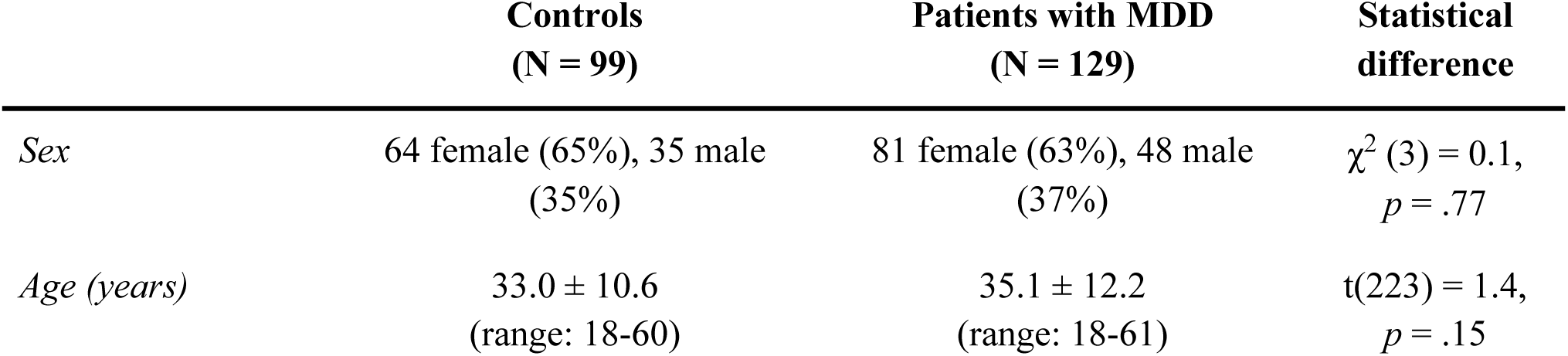
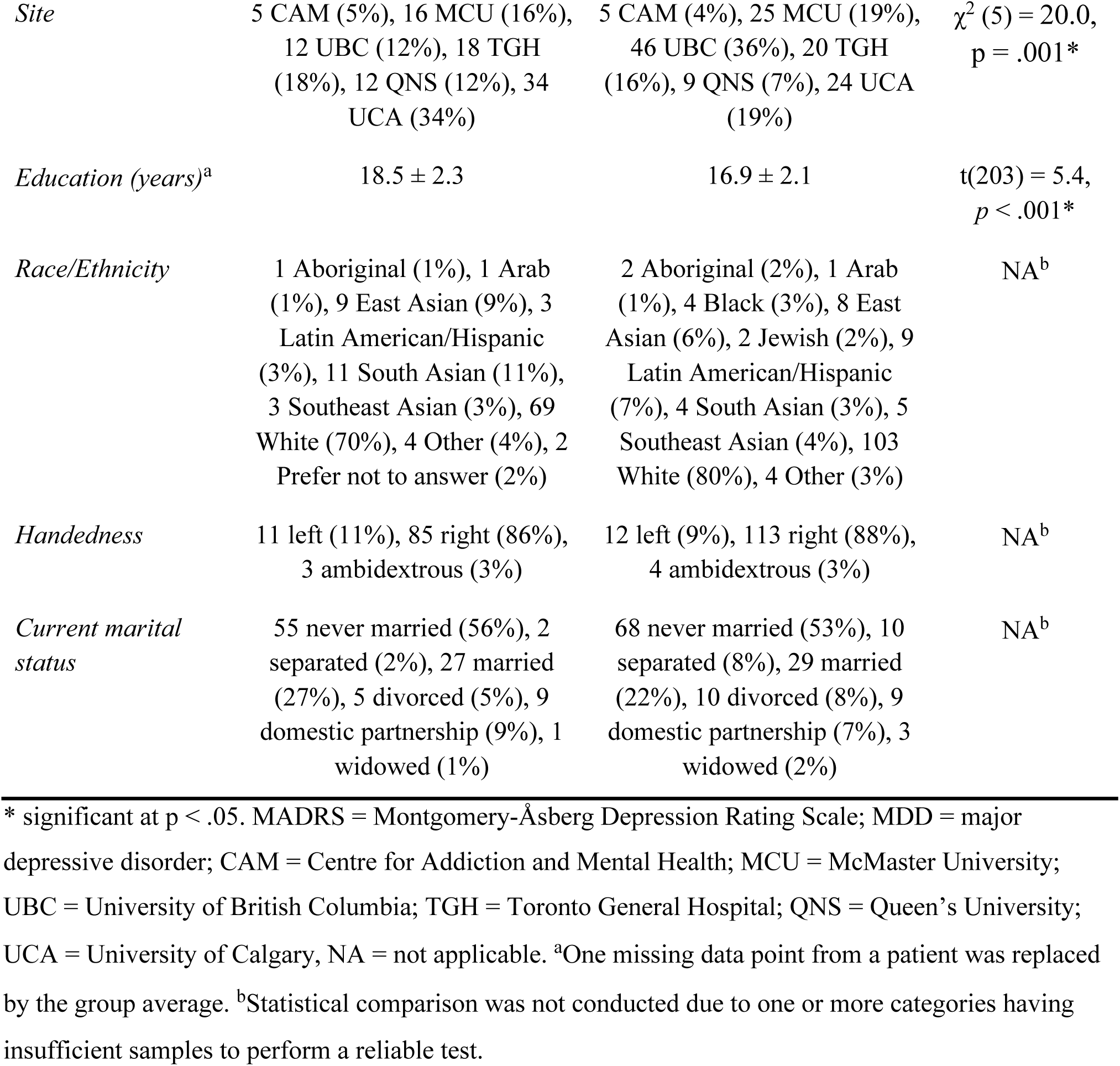
Characteristics of Controls and Patients with MDD (Mean ± SD), and their Statistical Differences

**Table 2.** Characteristics of Early Remitters, Late Remitters and Non-Remitters to Antidepressant Pharmacotherapy (Mean ± SD), and their Statistical Differences

|  | Early remitters<br>(N = 36) | Late remitters<br>(N = 46) | Non-remitters<br>(N = 47) | Statistical<br>difference |
| --- | --- | --- | --- | --- |
| *Sex* | 23 female (64%),<br>13 male (36%) | 30 female (65%),<br>16 male (35%) | 28 female (60%),<br>19 male (40%) | $\chi^2 (3) = 0.3$,<br>$p = .84$ |
| *Age (years)* | 34.6 ± 11.7 | 34.3 ± 12.1 | 36.4 ± 12.9 | $F(2, 126) = 0.4$, |

| | (range: 21-59) | (range: 18-59) | (range: 18-61) | $p = .67$ |
| --- | --- | --- | --- | --- |
| *Site* | 1 CAM (3%), 8 MCU (22%), 16 UBC (44%), 3 TGH (8%), 2 QNS (6%), 6 UCA (2%) | 1 CAM (2%), 11 MCU (24%), 12 UBC (26%), 8 TGH (17%), 4 QNS (9%), 10 UCA (22%) | 3 CAM (6%), 6 MCU (13%), 18 UBC (38%), 9 TGH (19%), 3 QNS (6%), 8 UCA 17% | NA<sup>b</sup> |
| *Education (years)<sup>a</sup>* | 17.1 ± 2.3 | 16.7 ± 2.1 | 16.9 ± 2.0 | F(2, 126) = 0.2, $p = .80$ |
| *Race/Ethnicity* | 1 Black (3%), 3 East Asian (8%), 2 Latin American/Hispanic (6%), 1 South Asian (3%), 1 Southeast Asian (3%), 28 White (78%), 2 Other (6%) | 1 Aboriginal (2%), 1 Arab (2%), 2 East Asian (4%), 2 Jewish (4%), 4 Latin American/Hispanic (9%), 1 South Asian (2%), 1 Southeast Asian (2%), 36 White (78%), 1 Other (2%) | 1 Aboriginal (2%), 3 Black (6%), 3 East Asian (6%), 3 Latin American/Hispanic (6%), 2 South Asian (4%), 3 Southeast Asian (6%), 39 White (83%), 1 Other (2%) | NA<sup>b</sup> |
| *Handedness* | 3 left (8%), 33 right (92%) | 3 left (7%), 42 right (91%), 1 ambidextrous (2%) | 6 left (13%), 38 right (81%), 3 ambidextrous (6%) | NA<sup>b</sup> |
| *Current marital status* | 18 never married (50%), 3 separated (8%), 9 married (25%), 3 divorced (8%), 2 domestic partnership (6%), 1 widowed (3%) | 28 never married (61%), 4 separated (9%), 8 married (17%), 1 divorced (2%), 3 domestic partnership (7%), 2 widowed (4%) | 22 never married (47%), 3 separated (6%), 12 married (26%), 6 divorced (13%), 4 domestic partnership (9%) | NA<sup>b</sup> |
| *Duration of current episode (months)<sup>a</sup>* | 25.5 ± 29.6 (range: 2-131) | 23.0 ± 30.9 (range: 1-130) | 37.2 ± 40.4 (range: 2-151) | F(2, 126) = 2.2, $p = .11$ |
| *Number of previous MDD episodes<sup>a</sup>* | 3.3 ± 3.0 (range: 0-13) | 3.1 ± 3.1 (range: 0-15) | 3.3 ± 4.1 (range: 0-20) | F(2, 126) = 0.04, $p = .96$ |
| *Baseline MADRS score* | 26.6 ± 4.3 | 30.9 ± 5.3 | 30.4 ± 5.6 | F(2, 126) = 7.9, $p = .001^*$ |
| *MADRS after 8 weeks of treatment* | 4.8 ± 3.1 | 18.5 ± 5.6 | 23.4 ± 8.3 | F(2, 126) = 94.4,<br>*p* < .001* |
| *MADRS after 16 weeks of treatment* | 3.2 ± 3.0 | 5.5 ± 3.0 | 19.3 ± 6.3 | F(2, 126) = 163.8,<br>*p* < .001* |
| *Treatment regimen* | 36 ESC (100%) | 13 ESC; 33<br>ESC+ARI | 5 ESC; 42<br>ESC+ARI | $\chi^2$ (2) = 72.3,<br>*p* < .001* |

\* significant at $p < .05$ . MADRS = Montgomery-Åsberg Depression Rating Scale; MDD = major depressive disorder; CAM = Centre for Mental Health and Addiction; MCU = McMaster University; UBC = University of British Columbia; TGH = Toronto General Hospital; QNS = Queen's University; UCA = University of Calgary; ESC = escitalopram; ARI = aripiprazole; NA = not available. <sup>a</sup>Missing data points (max. 3 per group) from individual participants were replaced by the group average.
<sup>b</sup>Statistical comparison was not conducted due to one or more categories having insufficient samples to perform a reliable test.

### MRI data acquisition and preprocessing

The structural and functional MRI scans used in this study were collected before the start of treatment. Resting-state fMRI scans were 10 minutes long, during which patients were asked to relax, have their eyes open, and focus on a fixation cross. A subgroup of 104 participants (64 patients and 40 controls) also performed an affective go/no-go task, lasting about 10 minutes. During the task, participants made go/no-go responses to stimuli presented on images containing affective content (see [27] for a detailed description of the task). Task instructions, stimulus presentation and task support materials were standardized across sites to ensure consistency. E-Prime version 2.0 (Psychology Software Tools, Sharpsburg, PA, USA) was used to record behavioural data. In the current study, go/no-go task data were only used for regions of interest (ROI) or “seed” selection (see *Seed selection* below).

Detailed information on the CAN-BIND MRI protocols can be found in MacQueen et al. [28]. Four different models of MRI scanners, all using 3.0 Tesla MRI systems with multicoil phased-array head coils, were used for data collection (Discovery MR750 3.0T, GE Healthcare; Signa HDxt 3.0T, GE Healthcare; MAGNETOM TrioTim, Siemens Healthcare; Achieva 3.0T, Philips Healthcare). To ensure data could be validly aggregated across sites, thorough quality control and standardization procedures were applied (see [17] for details). The anatomical scans were obtained using a whole-brain T1-weighted ‘magnetization prepared gradient echo sequence with the following parameters: voxel dimensions (in mm) = 1×1×1, echo time (TE) = 2.2-2.9ms, repetition time (TR) = 6.2-1900ms, inversion time (TI) = 450-950 ms, flip angle = 8 or 15°, field of view (FOV) = 240-256mm, matrix =240×240 or 256×256, number of slices = 176-192. The large range in TR was due to Siemens scanners reporting very different values for their proprietary sequence compared to GE and Philips, however image parameters were visually optimized to produce similar contrast across scanners. To aid the confirmation of participant orientation, a small Vitamin E capsule was used as a stereotactic marker by placing it at the right temple during recording. Functional scans were acquired with a whole-brain T2*-sensitive blood oxygenation level dependent (BOLD) echo planar imaging (EPI) series, with the following parameters: voxel dimensions (in mm) = 4×4×4, echo time (TE) = 25 or 30ms, repetition time (TR) = 2s, flip angle = 75° or 90°, field of view (FOV) = 256mm, matrix =64×64, number of slices = 34-40.

The resting state fMRI data were preprocessed with the OPPNI pipeline ([29, 30]; software available at https://github.com/raamana/oppni) using the following steps: 1) the volume with the least amount of head displacement was determined using a principal component analysis (PCA) and all volumes were registered to this volume with rigid-body motion correction (MOTCOR) via AFNI’s 3dvolreg; 2) significant outlier volumes were identified, removed and replaced by interpolated values using neighbouring volumes through censoring (CENSOR) as implemented in [31] (software available at: nitrc.org/projects/spikecor_fmri); 3) slice-timing correction (TIMECOR) was performed with Fourier interpolation via AFNI’s 3dTshift; 4) spatial smoothing across MRI scanners at different sites was matched using the 3dBlurToFWHM module in AFNI to smooth the fMRI images to the smoothness level of FWHM=6mm in three directions (x,y,z); 5) AFNI’s 3dAutomask algorithm was used to obtain a binary mask excluding non-brain voxels using default parameter settings, the resultant mask was applied to all EPI volumes prior to subsequent pipeline steps; 6) neuronal tissue masking was performed by estimating a probabilistic mask to reduce the variance contribution of non-neuronal tissues in the brain (macro-vasculature, ventricles) using the first part of the PHYCAA+ algorithm to estimate task-run and subject-specific neural tissue masks ([32]; software available at nitrc.org/projects/phycaa_plus); 7) several nuisance regressors (low frequency temporal trends, head motion effects and global signal modulations) were calculated and then regressed-out from the data concurrently via multiple linear regression [33–35]; 8) physiological noise components were estimated and removed through data-driven physiological correction (PHYPLUS) using the second part of the data-driven PHYCAA+ algorithm ([32]; software at nitrc.org/projects/phycaa_plus); 9) low-pass filtering (LOWPASS) was carried out using a linear filter to remove BOLD frequencies above 0.10 Hz; 10) spatial normalization to a structural template (sNORM) was carried out with all scans aligned to the MNI152 template (4mm resolution) using two transformations (fMRISubj -> MRISubj, and MRISubj→MNITemp, combined into one aggregated transform) via FSL’s FMRIB’s Linear Image Registration Tool (FLIRT) module.

The same steps were performed for the task fMRI data, except that no corrections were applied for global signal modulations, physiological noise components or higher frequency BOLD signals. After the OPPNI pipeline, the EPInorm strategy was used for additional spatial normalization on both the resting-state and task fMRI data, as this was shown to consistently reduce variability across participants and lower estimates for co-registration distances among participants [36]. Briefly, the data were registered directly to a cohort-specific EPI template using an affine followed by a nonlinear transformation. Only voxels in the grey matter, as determined by a grey matter mask, were included in the analyses. To account for potential differences in signal quality and tissue coverage, voxels from the grey matter mask were excluded from analysis on the basis of signal-to-noise ratio (SNR, voxel-wise intensity mean over the imaging time-course, divided by the standard deviation) where voxels were excluded if the calculated SNR was less than 100 in at least 5% of participants [37]. In our analysis this resulted in the exclusion of 2227 of the 17965 voxels originally in the grey matter mask.

### Seed selection

Four seeds were selected for analysis from the anterior cingulate cortex (ACC), the posterior cingulate cortex (PCC), the insula and the dorsolateral prefrontal cortex (dlPFC), because of their prominent role in three important networks associated with MDD pathology and antidepressant treatment response, namely the anterior DMN, posterior DMN, SN and CCN, respectively [4, 6–8]. Two seeds were selected from the DMN, as the anterior and posterior sections of this network have been found to show distinct alterations in response to antidepressant treatment [38].

The selection of specific seed coordinates was based on analyses contrasting task and resting state BOLD activity in the subgroup of participants who performed the affective go/no-go task. As the affective go/no-go task has been found to increase activation in both the SN and the CCN compared to rest [39, 40], the voxels with the most stable increase in activity (as indicated by bootstrap ratios, see *Data analysis with PLS*) during the task within the insula and dlPFC were selected as seeds for the SN and CCN. In contrast, the voxels showing the most stable increase in activation during rest compared to the task within the ACC and PCC were selected as the seeds for the anterior and posterior DMN. The average activation of the seed voxels and their immediate neighbours was extracted and used to examine functional connectivity between these regions of interest (ROIs) and the rest of the brain through correlation analyses.

This procedure was conducted twice to make the seed locations as representative as possible of the overall networks for the participants included in each of our two series of analyses: first for controls and patients together, to select the seeds for the patient-control comparisons, and then for patients only, to select the seeds for the comparisons between early, late and non-remitters. MDD and remission status were not considered in these seed selection analyses (i.e. participants were grouped together), to prevent seed selection from being influenced by group effects. Of note, the selected seeds were highly similar in both cases, except for the dlPFC seed (MNI coordinates patients & controls: ACC: [0 48 -4]; PCC: [0 -60 28]; Insula: [-40 20 0]; dlPFC: [-48 16 32]; MNI coordinates patients only: ACC: [0 48 -4]; PCC: [0 -60 28]; Insula: [-40 20 4]; dlPFC: [-52 28 28]).

### Data analysis with partial least squares (PLS)

Statistical analysis of functional connectivity was performed using the PLS Graphic User Interface version 6.1311050 (Rotman Research Institute, Ontario, Canada, http://www.rotman-baycrest.on.ca/pls) in MATLAB 2012a (The MathWorks, Inc., Natick, Massachusetts). PLS is a multivariate statistical technique that enables the detection of group and/or condition differences in the association between variables of interest and whole-brain activity patterns [41]. By entering the average BOLD activity extracted from the ROIs as variables of interest, we were able to examine functional connectivity as quantified by the correlations between the average activity in the ROIs and all other voxels of the brain across participants. PLS analyses identify latent variables (LVs), i.e. connectivity patterns highlighting similarities and/or differences between groups (MDD & controls/ER, LR & NR), that explain the largest amount of variance in the data. Each LV is made up of three components. The first is the singular value, which represents the strength of the effect expressed in the LV. The second contains the condition loadings, which indicate the contrast between groups as highlighted by the LV. The third component holds the element loadings, which describe where in the brain the connectivity differences identified by the LV are expressed.

The statistical assessment of LVs is done at two levels. First, the significance of the overall pattern is examined through permutation testing. Briefly, the data are randomly shuffled between groups and PLS analysis is then performed on the shuffled data for each permutation. LVs are considered significant when the singular value is more extreme than 95% of the singular values obtained from the shuffled data. We performed 1000 permutations per analysis. Second, bootstrap resampling is used to determine the consistency of the identified spatial pattern across participants. In this process, PLS analyses are repeated with different subsamples, building a distribution of loadings for each element depending on which participants are included. These are then used to calculate bootstrap ratios (BSRs) by dividing the element loadings by the standard error of the distribution for each element. BSRs represent the stability of the spatial pattern showing the LV contrast, and are similar to z-scores in that absolute BSR values ≥ 2 correspond to a confidence interval of ∼95%. We performed bootstrap resampling 1000 times per analysis. As the element loadings are all calculated in one mathematical step, no correction for multiple comparisons is necessary (i.e. patterns of connectivity across the whole brain are tested at once instead of individual connections).

First, we ran four PLS analyses comparing controls and all patients (grouped together), one for each ROI, to replicate previously found connectivity alterations in MDD. Next, we performed four PLS analyses including only patients, now grouped according to remission status (ER, LR, and NR), one for each of the four ROIs. For each significant LV, the spatial pattern was examined both manually and using the automated anatomical labeling (AAL) atlas [42] provided with the Fieldtrip toolbox [43]. Specifically, BSR thresholds were increased until the 10-15 clusters (involving 10 voxels or more) with the highest BSRs remained, and the areas in these clusters are reported in the results section. Manual inspection of these clusters was done using the MNI2TAL software developed by BioImage Suit and the Talairach atlas [44]. In addition, the voxels in these clusters were automatically labeled using the AAL atlas. Voxels that did not receive a label and regions with fewer than 3 voxels were not included. As we were primarily interested in group differences, we only report significant LVs that distinguished groups.

## Results

### Participants

There were no significant differences between patients and controls in the demographic characteristics presented in **Table 1**, except for controls having more years of education and a difference in the number of controls and patients tested at the different sites (**Table 1**). The three patient groups only differed in terms of their scores on the MADRS. By definition, early remitters had lower scores after 8 weeks of treatment, and both early and late remitters had lower scores after 16 weeks of treatment compared to non-remitters (**Table 2**). However, early remitters also showed lower MADRS scores at the start of treatment (**Table 2**). This baseline difference was beyond our control because group assignment was based on MADRS scores after 8 or 16 weeks of treatment.

### Replication of patient - control differences

#### Anterior cingulate ROI

The PLS analysis comparing connectivity between the ACC ROI and the rest of the brain between patients with MDD and controls identified one significant LV (*p* = .010, percent crossblock covariance explained (PCCE) = 22.8%) that highlighted differences between patients and controls. In line with our expectations, the pattern of differences included stronger connectivity between the ACC ROI and DMN regions (vmPFC, left PCC & left angular gyrus), the SN (right insula & dorsal ACC) and parts of the CCN (dmPFC & bilateral dlPFC) in patients compared to controls (see **Figure 1A**). In addition, this pattern included weaker connectivity between the ACC ROI and other parts of the CNN (bilateral PPC), the left extrastriate cortex and right premotor cortex, and stronger connectivity between the ACC ROI and the bilateral caudate nuclei, orbitofrontal cortex, left middle temporal gyrus and middle cingulate cortex in patients compared to controls. The labels of all the regions included in the 10-15 most stable clusters in this contrast, as determined by bootstrapping (BSR| ≥ 2.1), were identified using the AAL, and are presented in **Table S1**.

**Figure 1.**
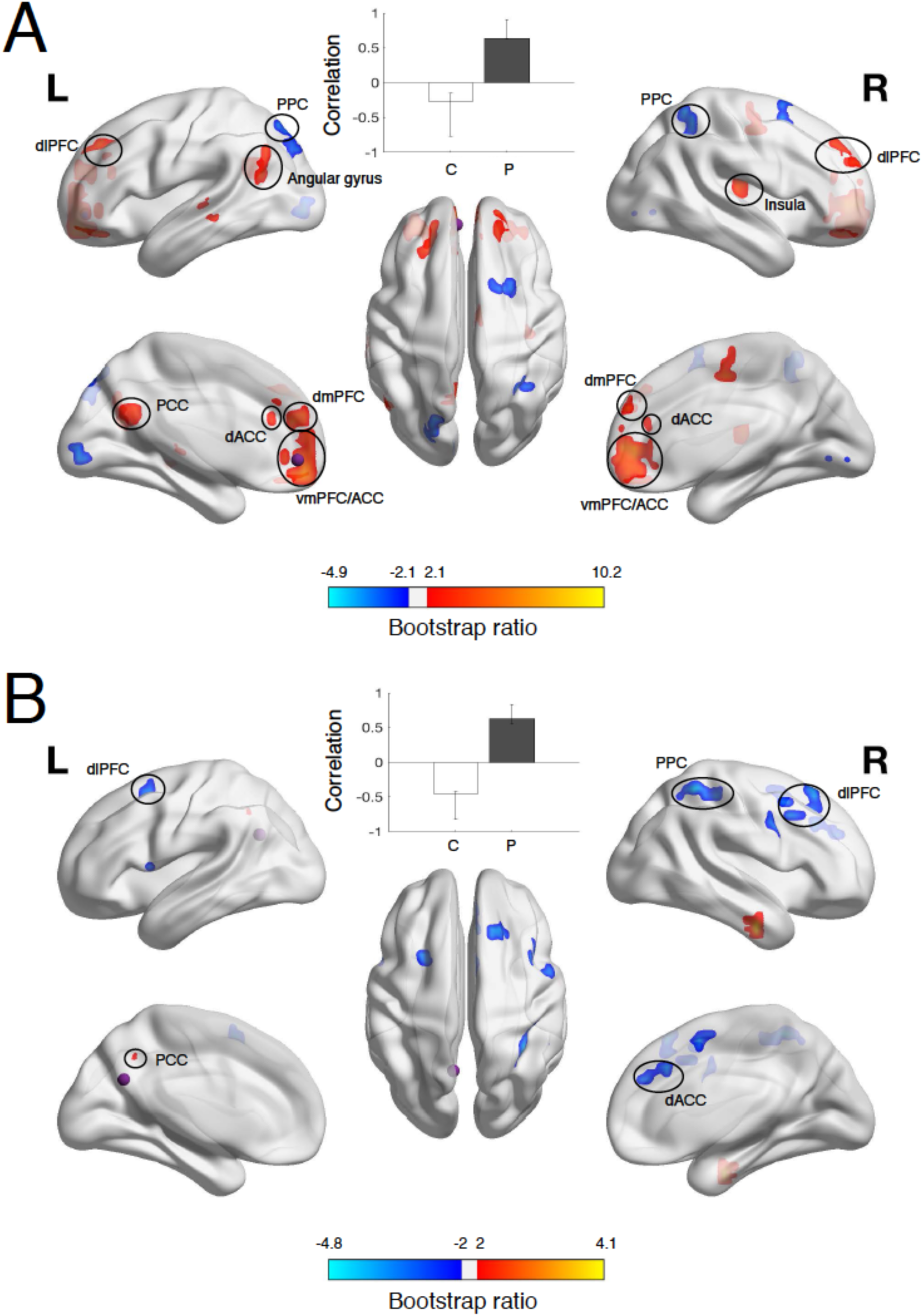
Results from PLS analyses comparing patients with MDD and controls in terms of A) connectivity between the ACC ROI (purple sphere) and all other voxels, and B) connectivity between the PCC ROI (purple sphere) and all other voxels, projected onto a smoothed cortical surface using BrainNet Viewer [45]. The correlation bar graph shows group-dependent differences in the correlation between the ROI voxels and the areas identified in the brain image. The error bars indicate the 95% confidence intervals derived from bootstrap estimation. The brain image illustrates the areas that expressed this contrast most stably across participants, as determined by bootstrapping. Only the 10-15 clusters (>10 voxels) with the highest bootstrap ratios (ACC: |BSR| ≥ 2.1; PCC: |BSR| ≥ 2.0) are presented. Cerebellar clusters are not illustrated, but they are listed in **table S1** (ACC) and **table S2** (PCC). Red/yellow clusters indicate stronger, while blue clusters indicate weaker connectivity in patients as compared to controls. C = controls, P = patients with major depressive disorder, dlPFC = dorsolateral prefrontal cortex, PPC = posterior parietal cortex, PCC = posterior cingulate cortex, dACC = dorsal anterior cingulate cortex, dmPFC = dorsomedial prefrontal cortex, vmPFC = ventromedial prefrontal cortex, ACC = anterior cingulate cortex.

#### Posterior cingulate ROI

The PLS analysis of connectivity between the PCC ROI and the rest of the brain in patients with MDD and controls did not identify significant LVs differentiating the two groups. However, one LV showing group differences in connectivity approached significance (p = .058, PCCE = 21.7%). As expected, this pattern included stronger connectivity between the PCC ROI and the precuneus (posterior DMN) and weaker connectivity between the PCC ROI and several CCN (bilateral dlPFC, right PPC) and SN (dorsal ACC) areas in patients compared to controls (see **Figure 1B**). In addition, this pattern included weaker connectivity between the PCC ROI and central motor areas (right SMA, bilateral premotor cortex), in patients compared to controls. The labels of all the regions included in the 10-15 most stable clusters, as determined by bootstrapping (|BSR| ≥ 2.0), were identified using the AAL and are presented in **Table S2**.

#### Insula ROI

The PLS analysis comparing patients with MDD and controls in connectivity between the insula ROI and the rest of the brain revealed one LV describing differences between patients and controls (p = .046, PCCE = 28.0%). As expected, the pattern of differences included weaker connectivity between the insula ROI and the PCC (posterior DMN) in patients compared to controls (see **Figure 2A**). Contrary to our expectations, we did not identify increased connectivity between the insula ROI and anterior DMN. In addition, the pattern included stronger connectivity between the insula ROI and the left dlPFC (from the CCN), occipital/posterior temporal regions in patients compared to controls. The labels of all the regions included in the 10-15 most stable clusters, as determined by bootstrapping (|BSR| ≥ 2.0), were identified using the AAL and are presented in **Table S3**.

**Figure 2.**
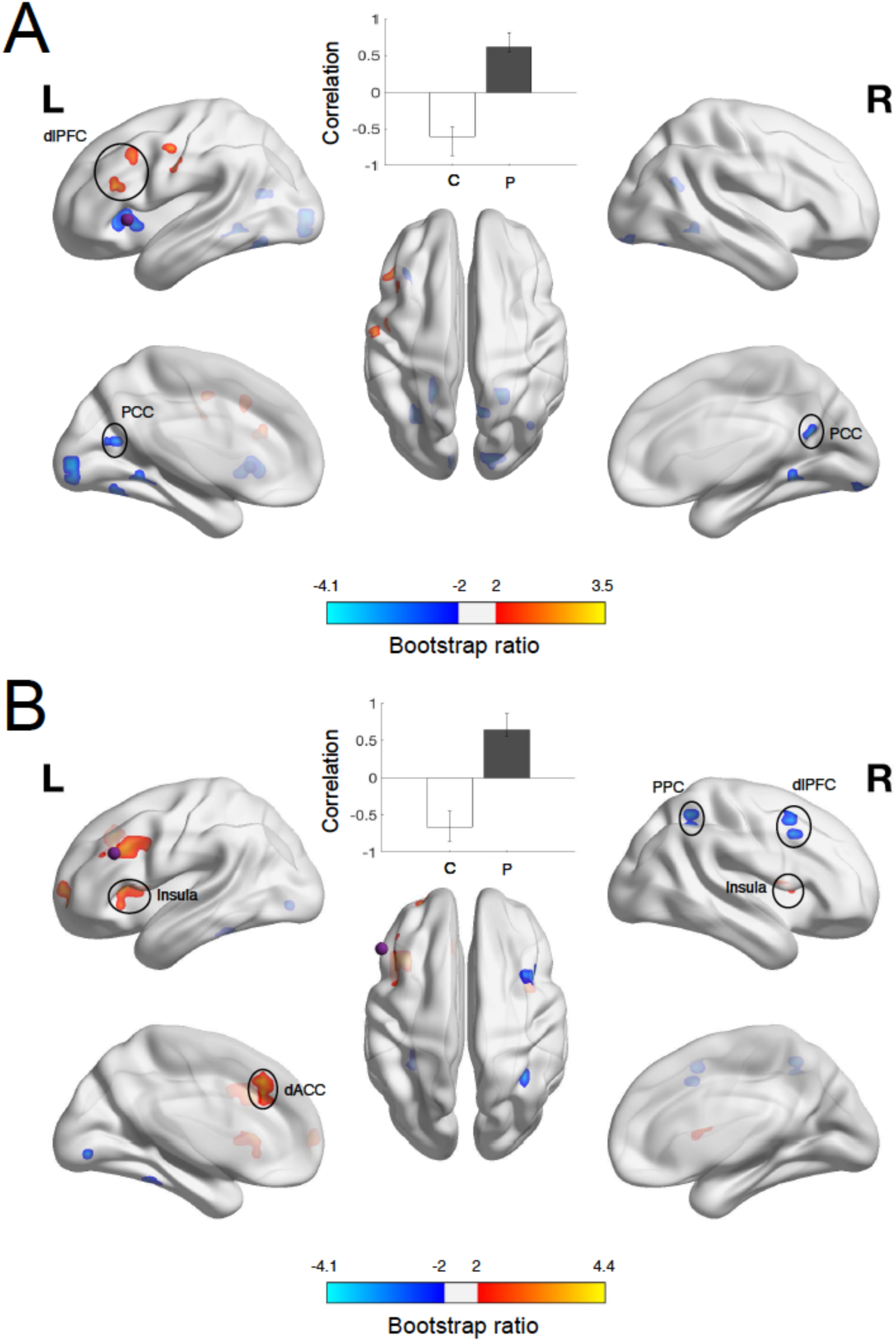
Results from PLS analyses comparing patients with MDD and controls in terms of A) connectivity between the insula ROI (purple sphere) and all other voxels, and B) connectivity between the dlPFC ROI (purple sphere) and all other voxels, projected onto a smoothed cortical surface using BrainNet Viewer [45]. The correlation bar graph shows group-dependent differences in the correlation between the ROI voxels and the areas identified in the brain image. The error bars indicate the 95% confidence intervals derived from bootstrap estimation. The brain image illustrates the areas that expressed this contrast most stably across participants, as determined by bootstrapping. Only the 10-15 clusters (>10 voxels) with the highest bootstrap ratios (insula: |BSR| ≥ 2.0; dlPFC: |BSR| ≥ 2.0) are presented. Cerebellar clusters are not illustrated, but they are listed in **table S3** (insula) and **table S4** (dlPFC). Red/yellow clusters indicate stronger, while blue clusters indicate weaker connectivity in patients compared to controls. C = controls, P = patients with major depressive disorder, dlPFC = dorsolateral prefrontal cortex, PPC = posterior parietal cortex, PCC = posterior cingulate cortex, dACC = dorsal anterior cingulate cortex.

#### Dorsolateral prefrontal cortex ROI

The PLS analysis of connectivity between the dlPFC ROI and the rest of the brain in patients and controls identified one LV revealing a pattern of group differences (p = .022, PCCE = 31.9%). In line with our expectations, this pattern included weaker connectivity between the dlPFC ROI and other CCN areas (right dlPFC, right PPC) in patients (see **Figure 2B**). At odds with our expectations, however, no connections between the dlPFC ROI and DMN regions were part of the pattern of differences. In addition, this pattern included stronger connectivity between the dlPFC ROI and SN regions (dorsal ACC & bilateral insulae), and the left anterior PFC and weaker connectivity the dlPFC ROI and visual processing areas (left extrastriate cortex, left fusiform gyrus) in patients compared to controls. The labels of all the regions included in the 10-15 most stable clusters, as determined by bootstrapping (|BSR| ≥ 2.0), were identified using the AAL and are presented in **Table S4**.

### Early, late and non-remitter comparison - anterior cingulate seed

The PLS analysis examining connectivity between the ACC ROI and all voxels in the brain across the patient groups (ER, LR, and NR) identified one significant LV (*p* = .008, PCCE = 31.4%) that differentiated between early remitters and the other two groups (see bar graph in **Figure 3**). This group specific contrast included stronger connectivity between the ACC ROI and the cerebellum, the PCC and the right occipital cortex (BA18), and weaker connectivity between the ACC ROI and the bilateral dlPFC, the medial anterior PFC, right subgenual ACC, right caudate nucleus, right superior temporal gyrus, left insula and the right extrastriate cortex (BA19) in early remitters compared to the other two groups (see **Figure 3**). The labels of all the regions included in the 10-15 clusters showing the most stable group differences, as determined by bootstrapping (|BSR| ≥ 2.4), were determined using the AAL atlas and are presented in **Table 3**.

**Table 3.**
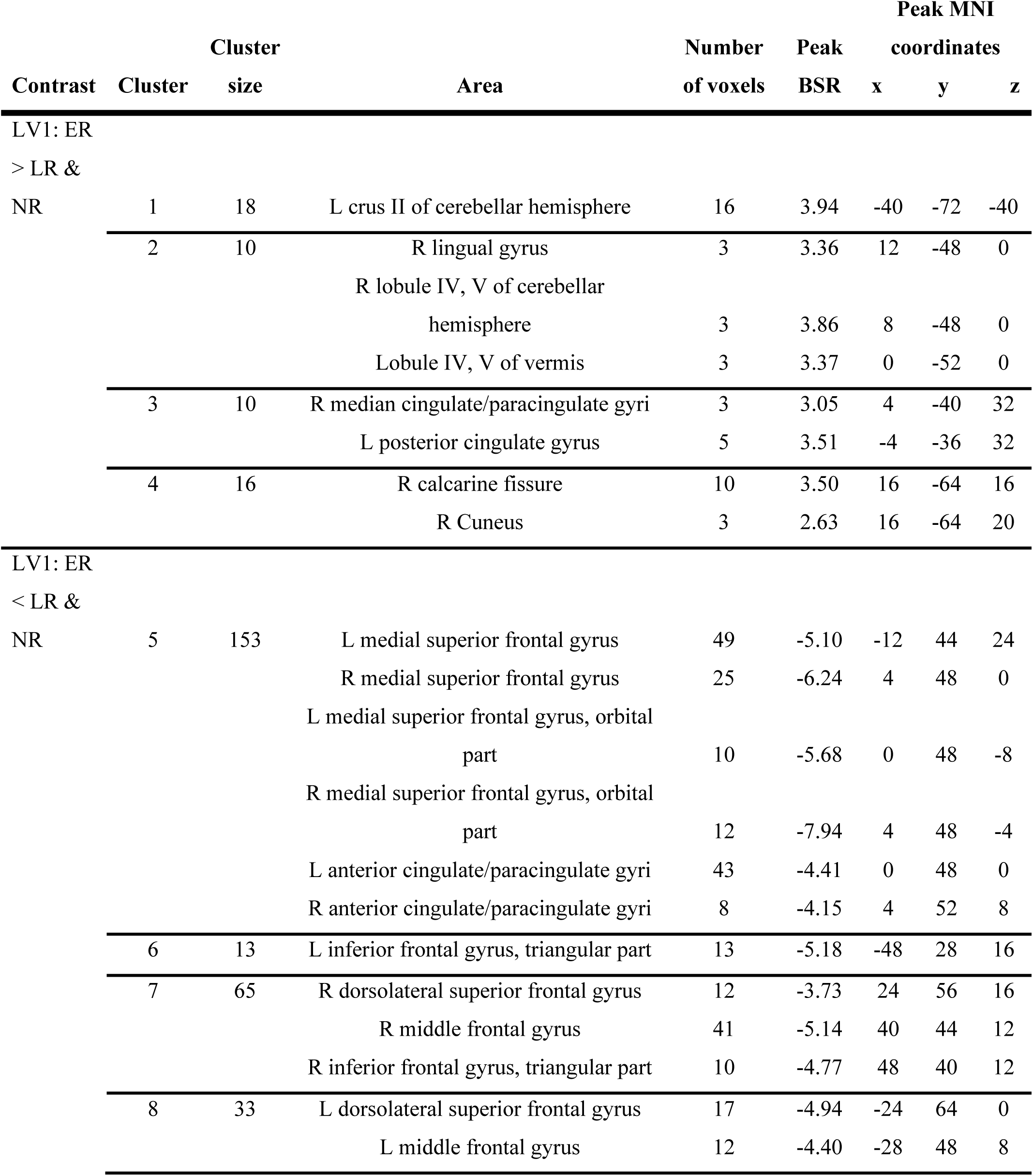
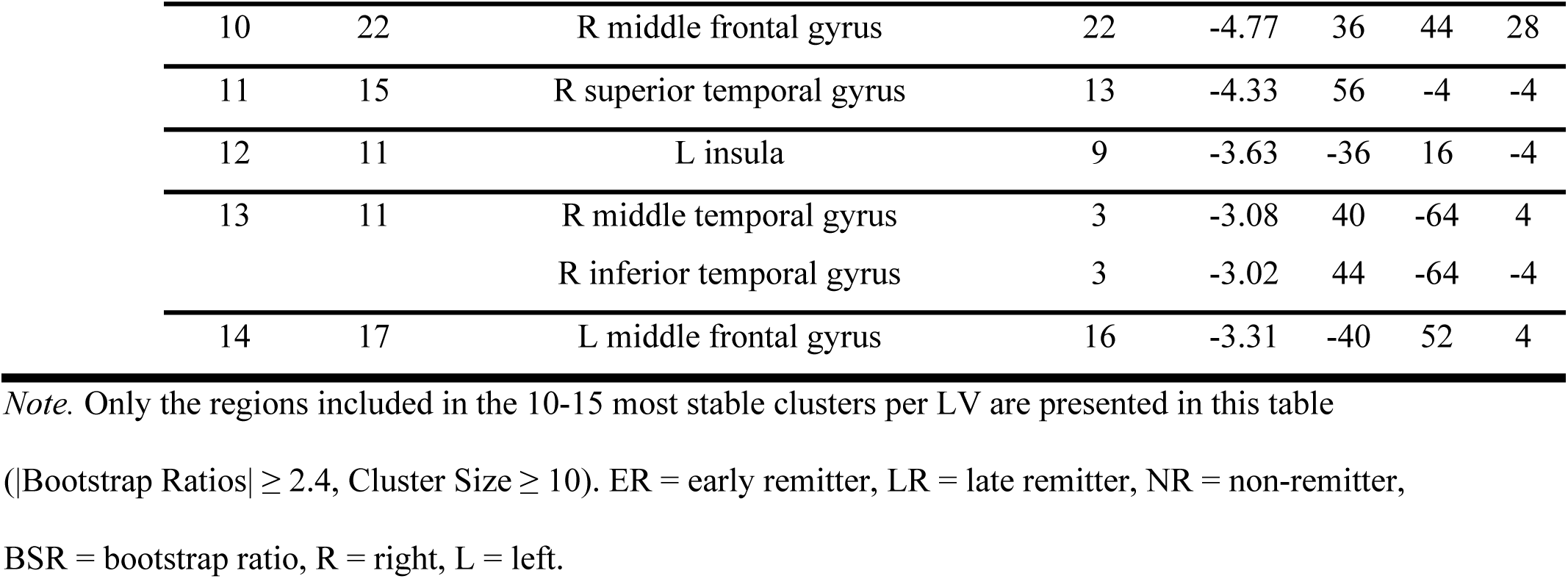
Regions Showing Distinct Connectivity with the Anterior Cingulate Cortex in Early, Late and Non-remitters Automatically Labeled Using the AAL Atlas

**Figure 3.**
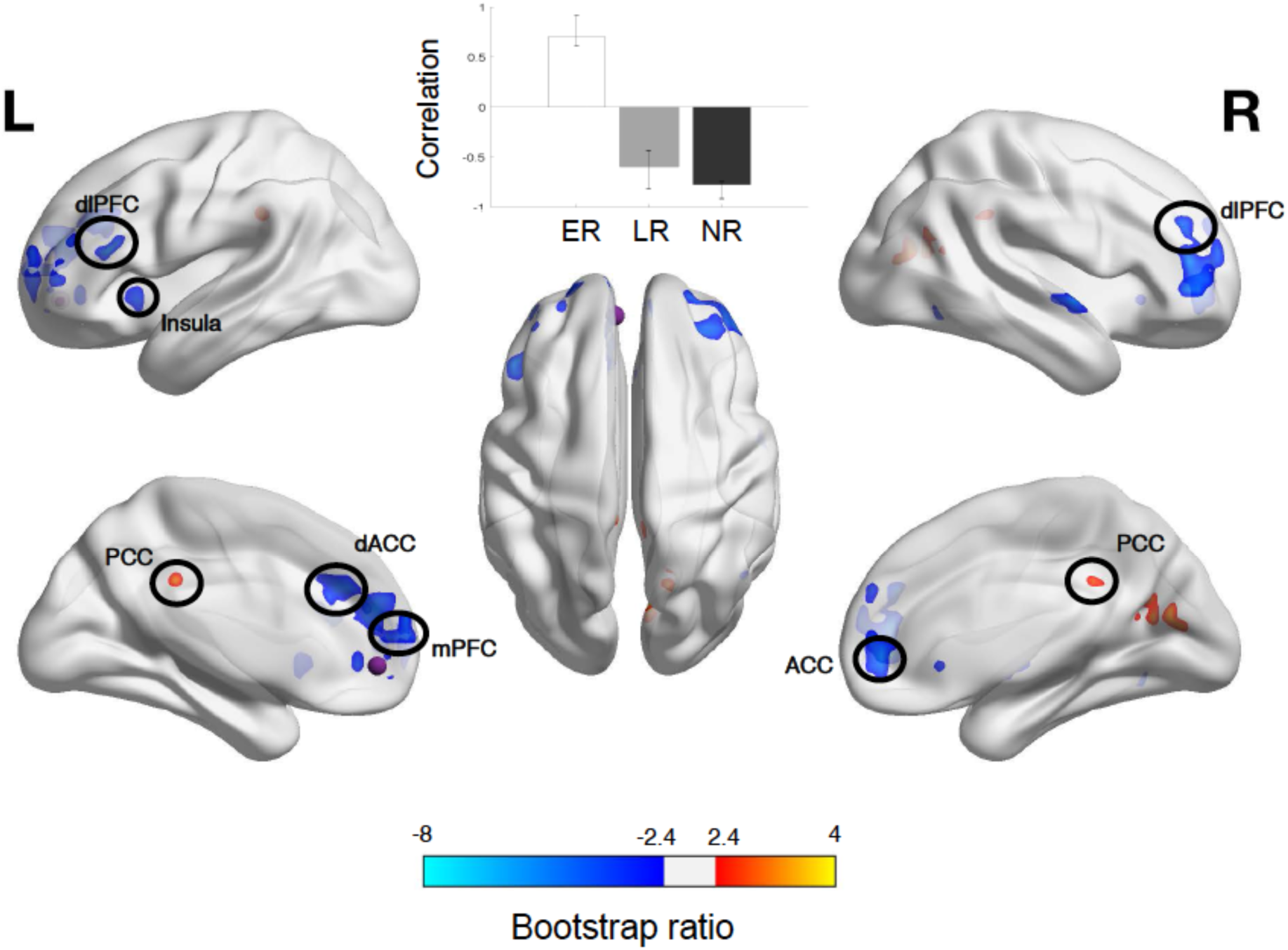
Results from PLS analysis comparing early, late and non-remitters in terms of connectivity between the ACC ROI (purple sphere) and all other voxels, projected onto a smoothed cortical surface using BrainNet Viewer [45]. The correlation bar graph shows group-dependent differences in the correlation between the ROI voxels and the areas identified in the brain image. The error bars indicate the 95% confidence intervals derived from bootstrap estimation. The brain image illustrates the areas that expressed this contrast most stably across participants, as determined by bootstrapping. Only the 10-15 clusters (>10 voxels) with the highest bootstrap ratios (|BSR| ≥ 2.4) are presented. Cerebellar clusters are not illustrated, but they are listed in **table 3**. Red/yellow clusters indicate stronger, while blue clusters indicate weaker connectivity in early compared to late and non-remitters. ER = early remitters, LR = late remitters, NR = non-remitters, dlPFC = dorsolateral prefrontal cortex, PCC = posterior cingulate cortex, dACC = dorsal anterior cingulate cortex, mPFC = medial prefrontal cortex, ACC = anterior cingulate cortex.

### Early, late and non-remitter comparison - posterior cingulate seed

The result of the PLS analysis investigating connectivity between the PCC ROI and all voxels in the brain identified one significant LV (p = .016, PCCE = 24.7%) which distinguished the three groups. Early and late remitters showed a pattern of connectivity that was most consistently expressed as stronger connectivity between the PCC ROI and the precuneus/right PCC, the cerebellum, the bilateral superior temporal gyrus, orbital PFC, dorsal and pregenual ACC and the left fusiform gyrus, and weaker connectivity between the PCC ROI and the right superior temporal gyrus compared to non-remitters. Importantly, early remitters exhibited this pattern of connectivity more strongly than late remitters (see bar graph in **Figure 4**). All regions identified in the 10-15 most stable clusters, as determined by bootstrapping (|BSR| > 2.3), were labeled using the AAL atlas and are listed in **Table 4**.

**Table 4.**
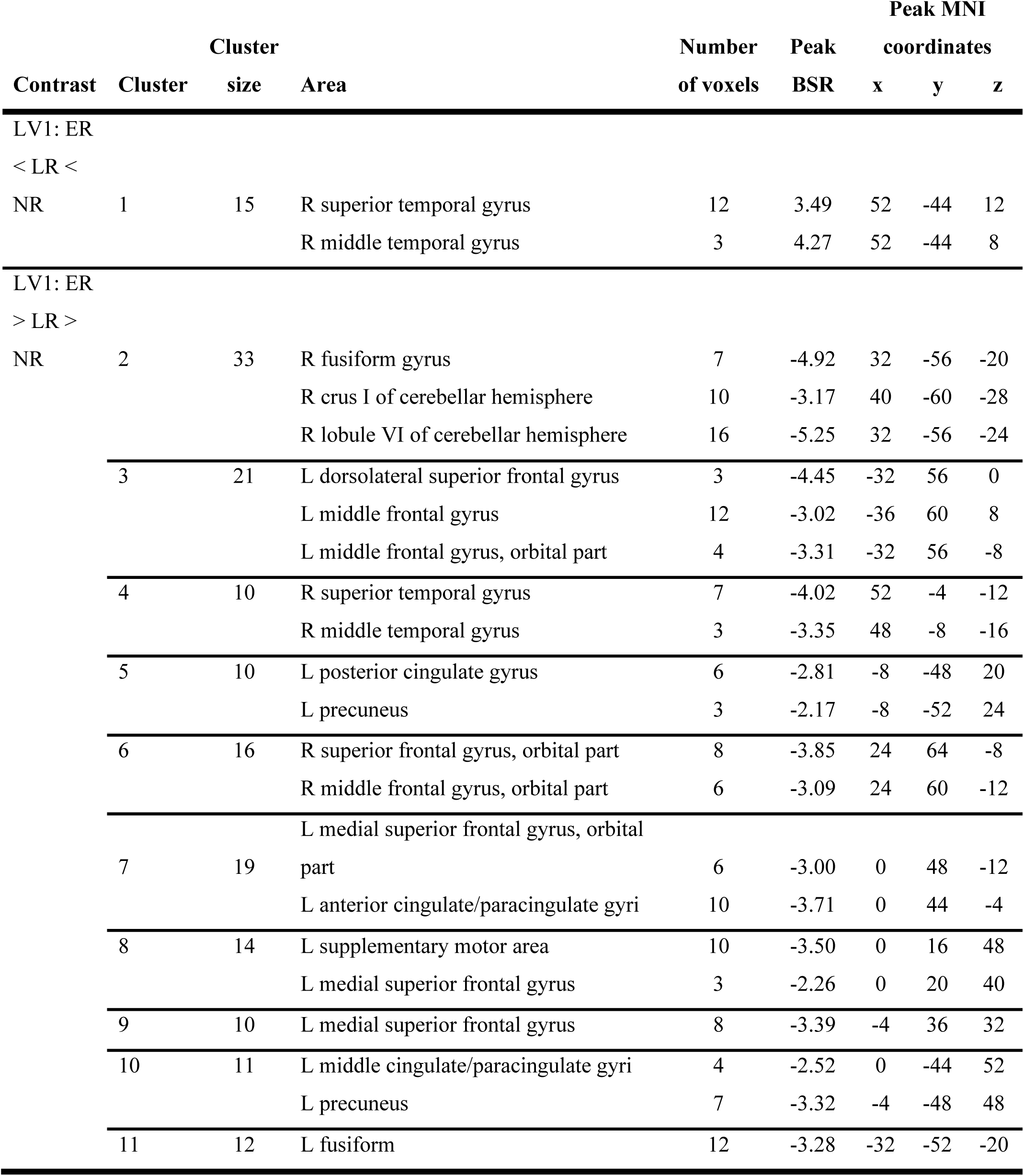
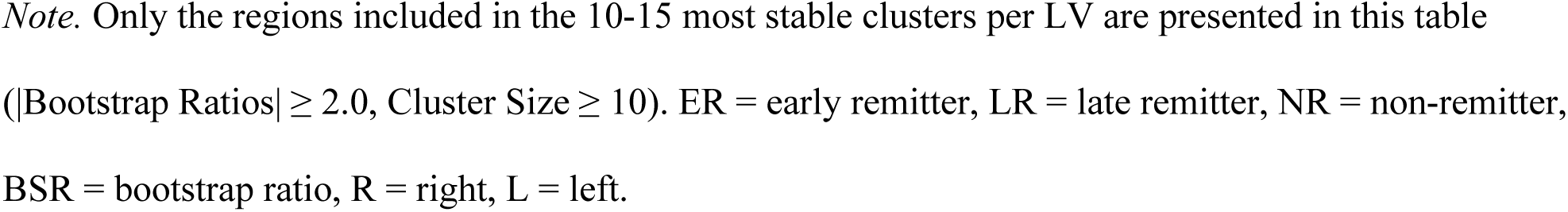
Regions Showing Distinct Connectivity with the Posterior Cingulate Cortex in Early, Late and Non-remitters Automatically Labeled Using the AAL Atlas

**Figure 4.**
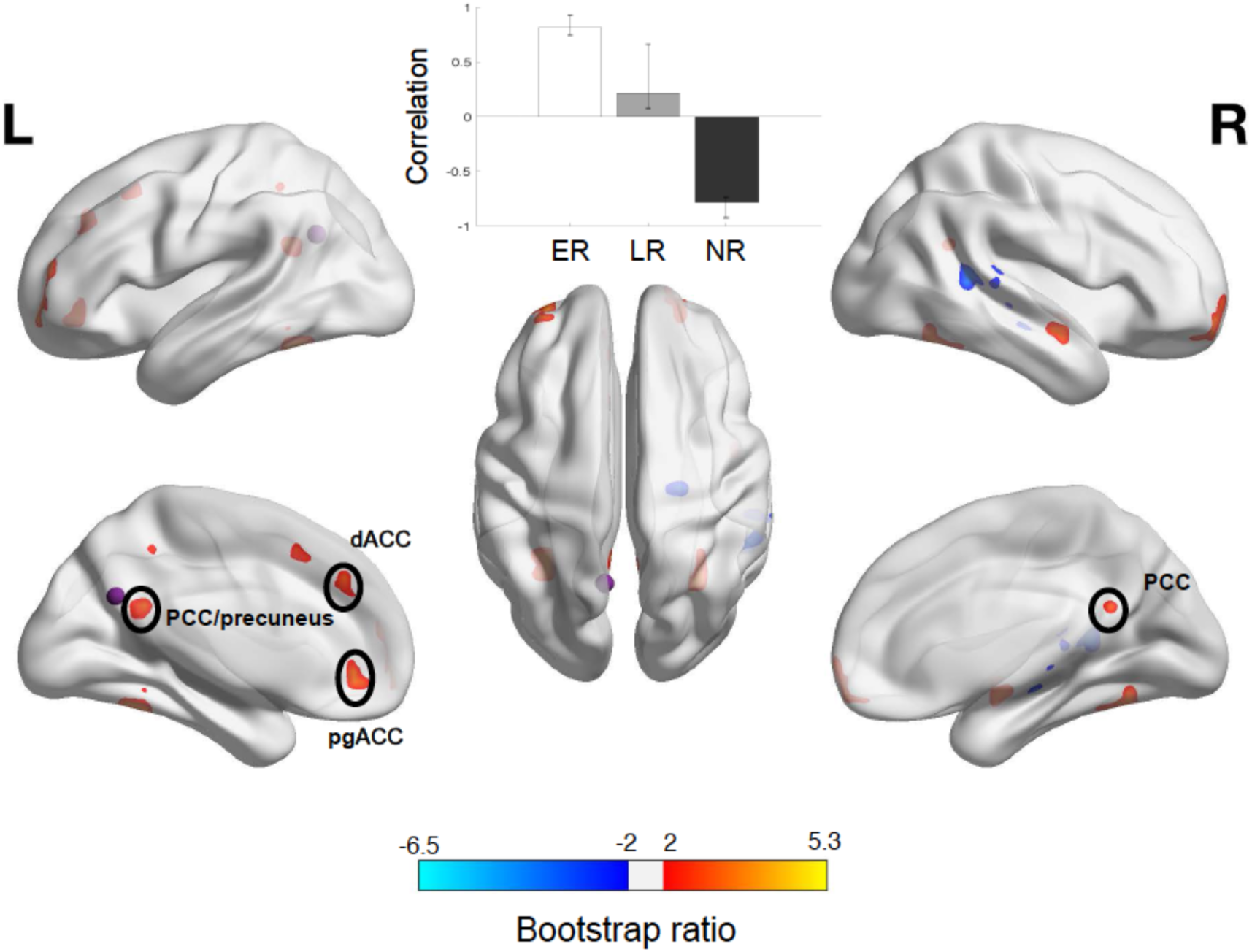
Results from PLS analysis comparing early, late and non-remitters in terms of connectivity between the PCC ROI (purple sphere) and all other voxels, projected onto a smoothed cortical surface using BrainNet Viewer [45]. The correlation bar graph shows group-dependent differences in the correlation between the ROI voxels and the areas identified in the brain image. The error bars indicate the 95% confidence intervals derived from bootstrap estimation. The brain image illustrates the areas that expressed this contrast most stably across participants, as determined by bootstrapping. Only the 10-15 clusters (>10 voxels) with the highest bootstrap ratios (|BSR| ≥ 2.4) are presented. Cerebellar clusters are not illustrated, but they are listed in **table 4**. Red/yellow clusters indicate stronger, while blue clusters indicate weaker connectivity in early remitters compared to late and non-remitters, as well as in late compared to non-remitters. ER = early remitters, LR = late remitters, NR = non-remitters, PCC = posterior cingulate cortex, dACC = dorsal anterior cingulate cortex, pgACC = pregenual anterior cingulate cortex.

### Early, late and non-remitter comparison - insula ROI

The PLS analysis of connectivity between the insula ROI and the rest of the brain in early, late and non-remitters identified two significant LVs (LV1: p = .005; PCCE = 27.8%, LV2: p = .044, PCCE = 23.1%) differentiating between patient groups. The first differentiated late and early remitters, with early remitters showing stronger connectivity between the insula ROI and the PCC/precuneus, bilateral fusiform gyrus, PPC, anterior and medial PFC, ACC, the left FEF, the left dlPFC and the right angular gyrus, and weaker connectivity between the insula ROI and the right insula, right inferior frontal gyrus, the cerebellum and the left putamen, as compared to late remitters (see **Figure 5**). All regions identified in the 10-15 most stable clusters, as determined by bootstrapping (|BSR| ≥ 2.4), were labeled using the AAL atlas and are listed in **Table 5**.

**Table 5.**
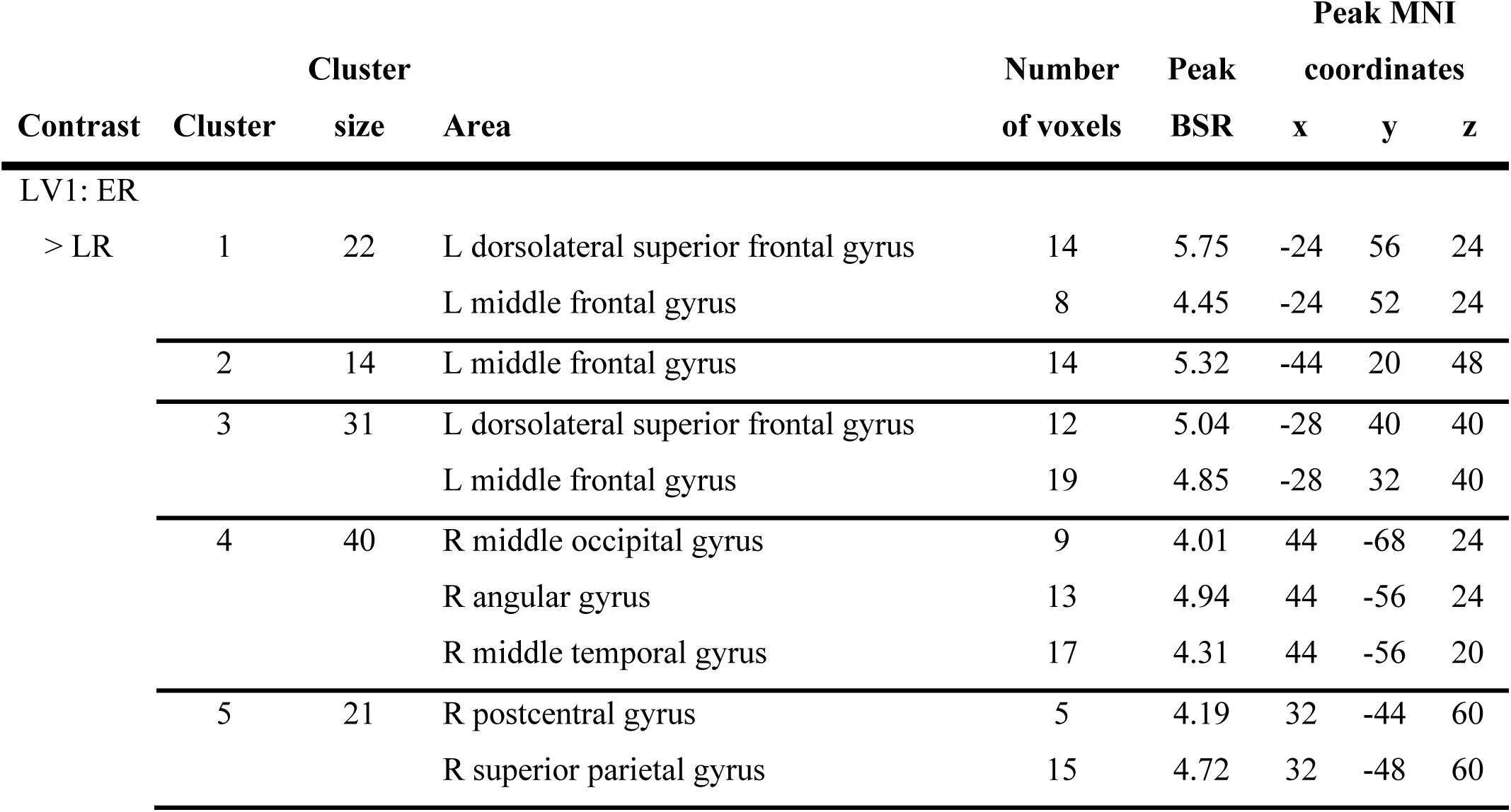
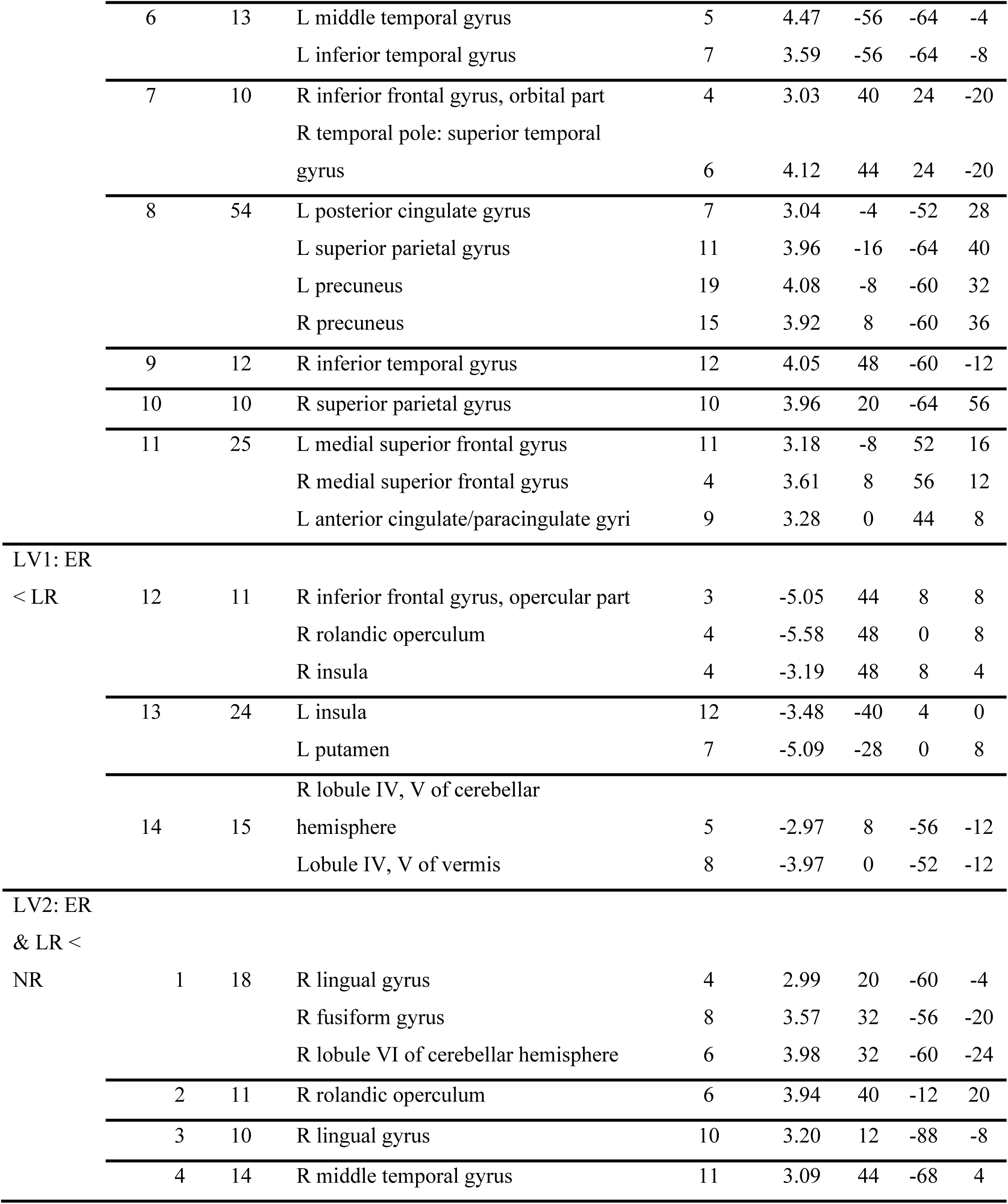
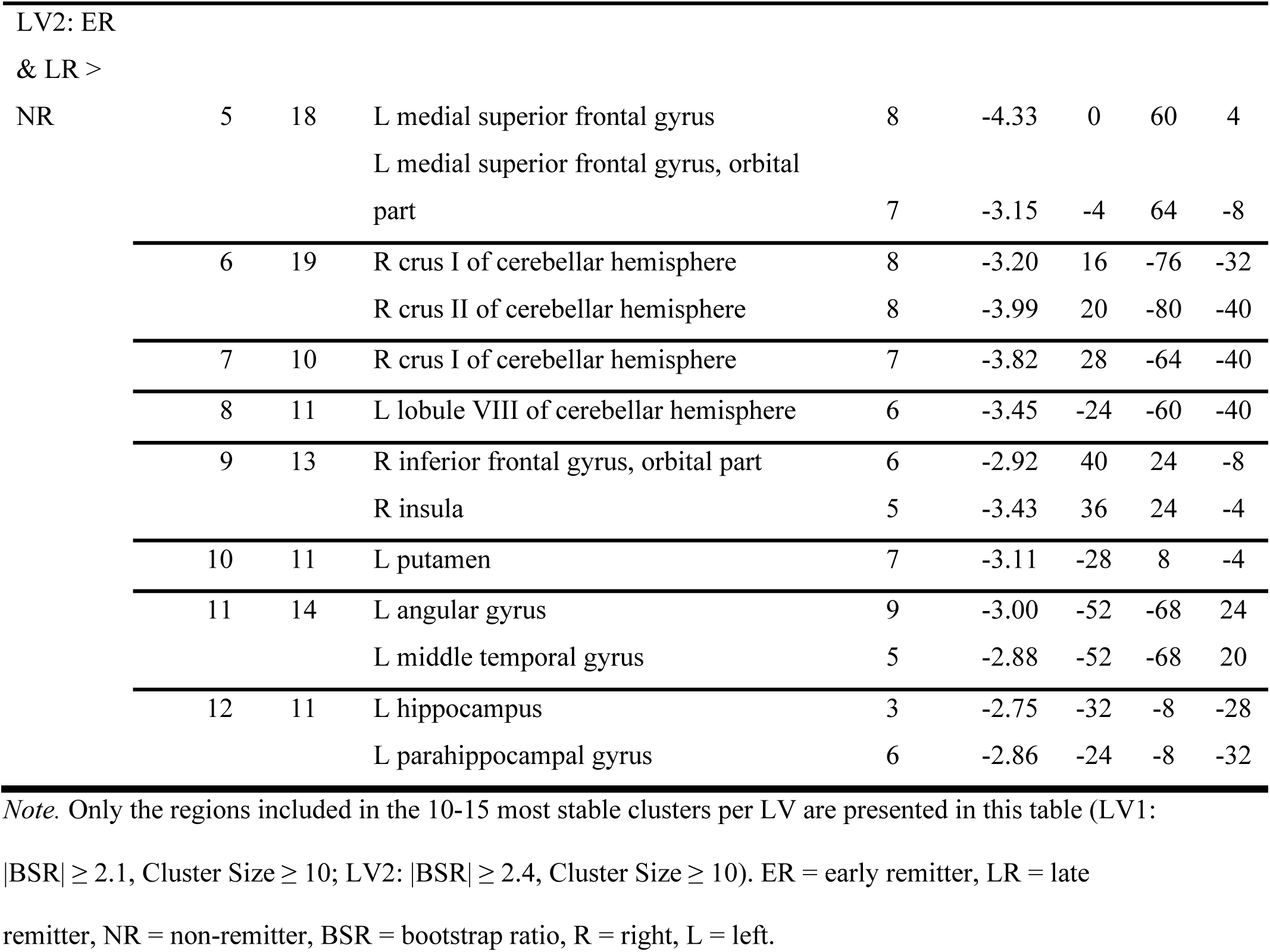
Regions Showing Distinct Connectivity with the Insula in Early, Late and Non-remitters, Automatically Labeled Using the AAL Atlas

**Figure 5.**
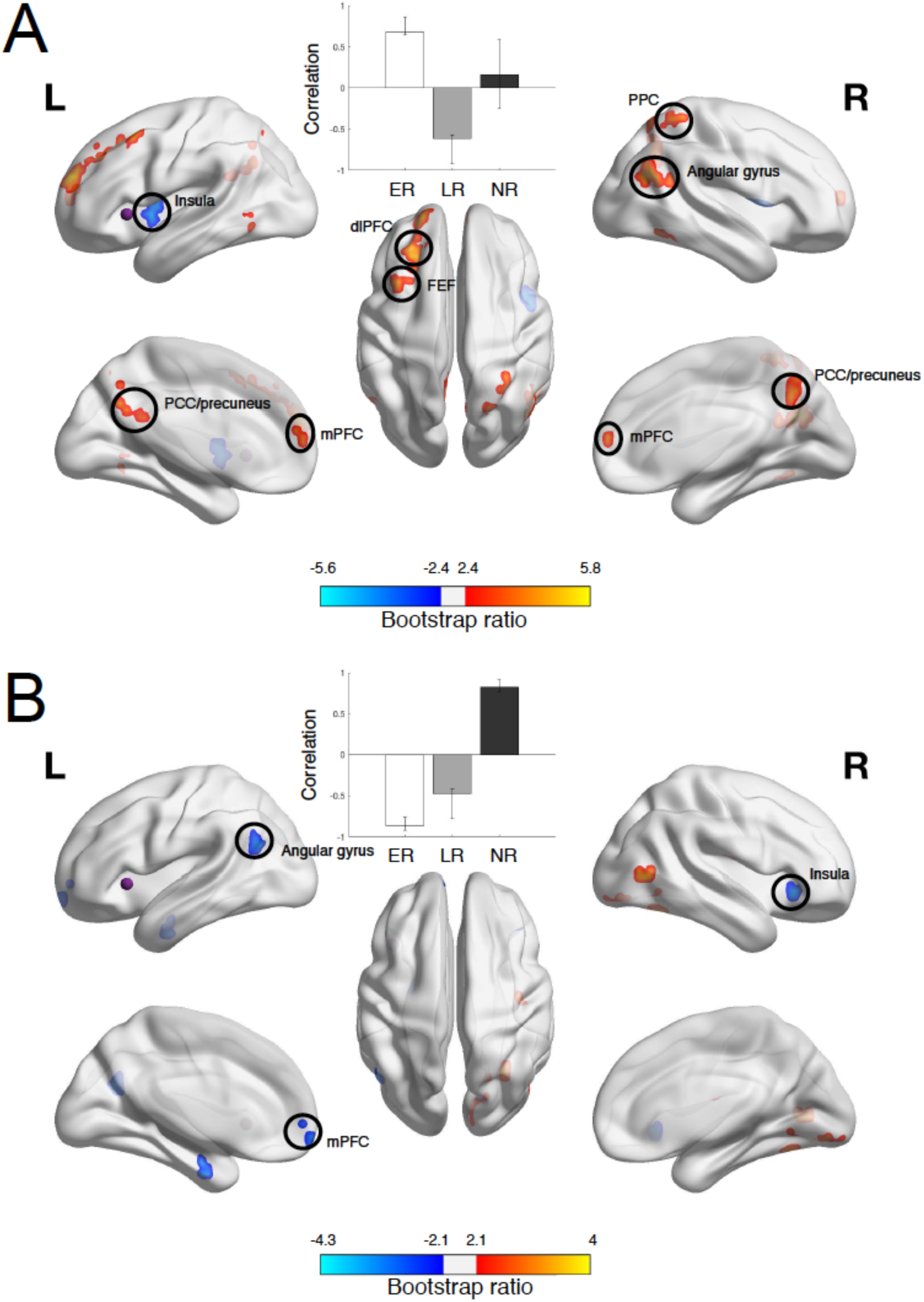
Results from PLS analysis comparing early, late and non-remitters in terms of connectivity between the insula ROI (purple sphere) and all other voxels, projected onto a smoothed cortical surface using BrainNet Viewer [45]. A) shows the contrast and spatial pattern identified in the first LV, B) contrast and spatial pattern identified for the second LV. The correlation bar graph shows group-dependent differences in the correlation between the ROI voxels and the areas identified in the brain image. The error bars indicate the 95% confidence intervals derived from bootstrap estimation. The brain image illustrates the areas that expressed this contrast most stably across participants, as determined by bootstrapping. Only the 10-15 clusters (>10 voxels) with the highest bootstrap ratios (|BSR| ≥ 2.4) are presented. Cerebellar clusters are not illustrated, but they are listed in **table 5**. In A) red/yellow clusters indicate stronger, while blue clusters indicate weaker connectivity in early remitters compared to late remitters. In B) red/yellow clusters indicate stronger, while blue clusters indicate weaker connectivity in non-remitters compared to early and late remitters. ER = early remitters, LR = late remitters, NR = non-remitters, dlPFC = dorsolateral prefrontal cortex, PPC = posterior parietal cortex, FEF = frontal eye fields, PCC = posterior cingulate cortex, mPFC = medial prefrontal cortex, LV = latent variable.

The second LV distinguished between remitters and non-remitters. Specifically, non-remitters showed stronger connectivity between the insula ROI and the cerebellum, right extrastriate cortex and right operculum, and weaker connectivity between the insula ROI and the cerebellum, right insula, left angular gyrus, putamen, mPFC and parahippocampal gyrus compared to both early and late remitters. All regions identified in the 10-15 most stable clusters, as determined by bootstrapping (|BSR| ≥ 2.1), were labeled using the AAL atlas and are listed in **Table 5**.

### Early, late and non-remitter comparison - dorsolateral prefrontal cortex ROI

The PLS analysis comparing early, late and non-remitters in connectivity between the dlPFC ROI and the rest of the brain did not identify any significant LVs highlighting differences between patient groups.

### Subsample replication

To assess the stability of our patient group comparisons, we ran additional PLS analyses on the largest single-site population of patients (16 ER, 12 LR, & 18 NR, from the UBC site). Specifically, we used non-rotated (i.e. hypothesis-driven) PLS to test whether the group differences we identified in the main analyses with the ACC, PCC and insula ROI, were present in this subsample (one each for the ACC and PCC, two for the insula ROI). In addition to inspecting the analysis results visually, we correlated the stable (|BSR| ≥ 2) element loadings of the main analyses with those of the subsample analyses to assess their similarity. As these were post-hoc analyses, we applied Bonferroni correction to the results. The results are summarized in **Table 6**. Two out of the four LVs tested were significant according to the Bonferroni corrected p-value (*p* < .013), one fell into the uncorrected window of significance (*p* = .043), and one showed a trend towards significance (*p* = .092). Correlations between stable element loadings ranged between .80 and .90.

**Table 6.**
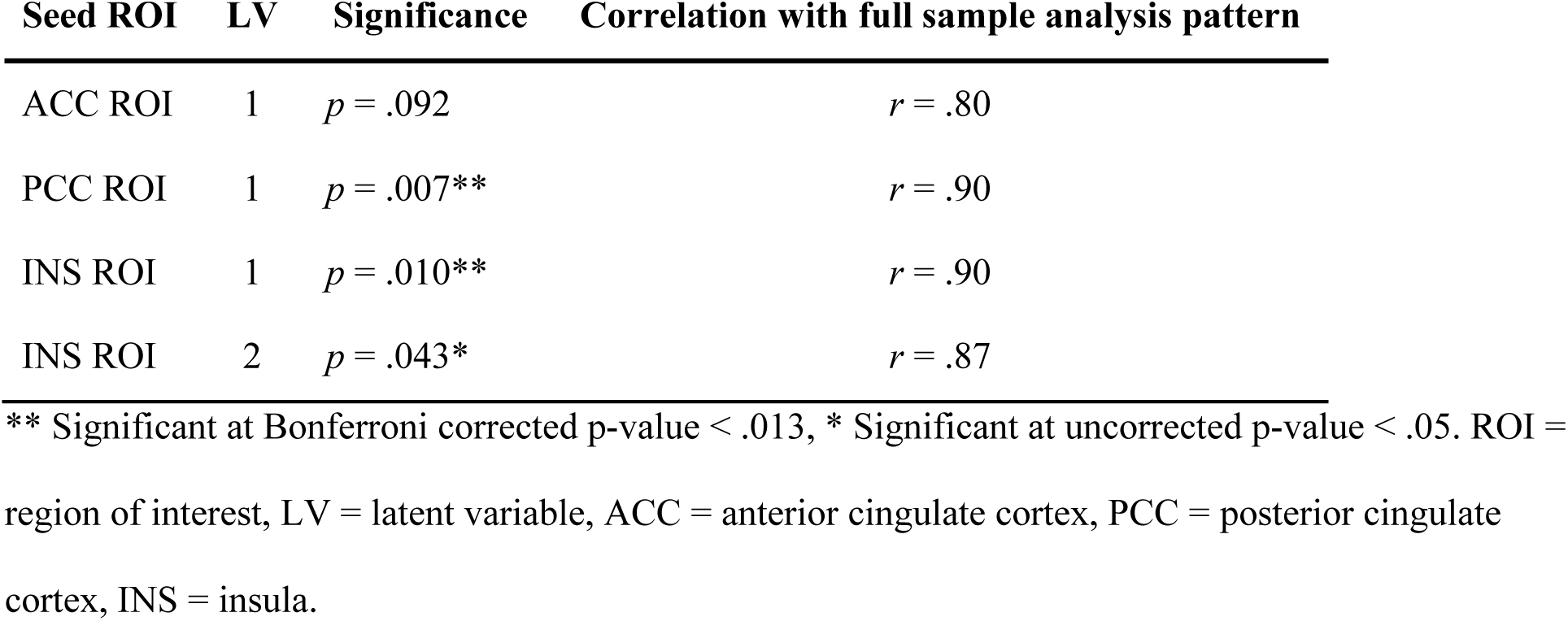
Significance and Correlation of Subsample Replication Analyses for Each Subsample and Seed ROI

| Seed ROI | LV | Significance | Correlation with full sample analysis pattern |
| --- | --- | --- | --- |
| ACC ROI | 1 | $p = .092$ | $r = .80$ |
| PCC ROI | 1 | $p = .007^{**}$ | $r = .90$ |
| INS ROI | 1 | $p = .010^{**}$ | $r = .90$ |
| INS ROI | 2 | $p = .043^{*}$ | $r = .87$ |

\*\* Significant at Bonferroni corrected p-value $< .013$ , \* Significant at uncorrected p-value $< .05$ . ROI = region of interest, LV = latent variable, ACC = anterior cingulate cortex, PCC = posterior cingulate cortex, INS = insula.

### Post-hoc analysis examining remission to escitalopram alone or combined escitalopram and aripiprazole

To test whether the differences observed in insula connectivity between early and late remitters (see *Early, late and non-remitter comparison - insula ROI)* were related to differences in the medication regimen received (see **Table 2**), we ran a PLS analysis examining differences in insula connectivity between remitters to escitalopram alone (N = 33) and remitters to combined escitalopram and aripiprazole (N = 49). This revealed one significant LV differentiating between the two groups (p < .001, PCCE = 37.4%), indicating that, before treatment, participants remitting to escitalopram alone most consistently showed stronger connectivity between the insula ROI and medial and anterior PFC areas and the dorsal ACC (see **Table S5** & **Figure S1**) compared to participants who remitted to combined escitalopram and aripiprazole.

## Discussion

We aimed to replicate and extend previous findings linking abnormal fMRI connectivity in three major resting state networks to the depressed state and antidepressant treatment outcomes using a large, well-characterized, multi-site sample. Overall, we were able to replicate the more robust differences between patients with MDD and controls highlighted in previous reviews [4, 5]. In addition, we identified differences in connectivity within and between these same networks within the patient population. ACC connectivity differentiated early from late and non-remitters, PCC connectivity distinguished early, late and non-remitters, and insula connectivity revealed both early/late remitter and remitter/non-remitter contrasts. Importantly, these differentiating connectivity patterns held in a single-site subsample, indicating a degree of stability of these group differences. As we looked at the relationship between connectivity before the start of treatment and remission status after 16 weeks of pharmacotherapy, these patterns might be useful in predicting antidepressant treatment outcomes.

In line with previous literature [4, 5], we found stronger connectivity within the DMN, most notably in the anterior subnetwork, and between the anterior DMN and the CCN and SN, and weaker connectivity within the CCN in patients as compared to controls. We also found weaker connectivity between the posterior DMN and the CCN and SN in patients, but this contrast only approached significance. Interestingly, altered connectivity between the anterior DMN and the SN, and the DMN and the CCN, was only apparent when using DMN regions as seeds (ACC & PCC ROIs). This could indicate that DMN abnormalities are stronger and thus most useful in distinguishing patients from controls. While the DMN certainly takes a prominent place in the literature on brain network alterations related to MDD, so do the two other networks investigated in this project. An alternative interpretation might be that DMN effects were stronger because we recorded data during resting state, when the DMN is typically active, while both the CCN and SN are typically more active during tasks [46].

In addition to these expected effects, the connectivity patterns differentiating patients and controls included connections that have not been highlighted in previous reviews. Most interestingly, we identified stronger connectivity between CCN and SN regions in patients using both the CCN and SN seeds (dlPFC and insula, respectively). Few studies have found altered connectivity between the CCN and SN directly (e.g. [47]). However, these two networks are involved in emotion processing and mood regulation, which have been found to be abnormal in patients with MDD (e.g. [48]), therefore our finding of aberrant connectivity between them is not surprising. Mulders et al. [4] mention that the SN is less well-defined during resting state (in comparison to the DMN and CCN), so effects might be harder to detect for this network, especially with small samples. Our finding of increased connectivity between the SN and CCN in a larger sample, together with the potential functional relevance of these connections, indicates that SN-CCN connectivity should be studied in more detail in future research. In addition, several regions outside of our networks of interest were highlighted by our analyses, including areas related to visual and motor processing. While generally given little attention, such regions have been observed in previous studies examining differences between patients with MDD and controls [4, 5], indicating that network alterations associated with MDD are more widespread than is generally highlighted.

Nonetheless, the three selected networks revealed important baseline connectivity differences between patients who reached remission early (within 8 weeks of treatment) or late (within 16 weeks), or who did not reach remission over the course of this study. Early remitters displayed weaker connectivity within the anterior DMN, and between the ACC seed and regions from the CCN and SN compared to both late and non-remitters. The ACC has been proposed to play a central role in MDD pathology [49–51]. While the subgenual part of the ACC is most often highlighted, the pregenual ACC, where our seed was located, has also been found to be part of the altered DMN in MDD and associated with treatment effects [52, 53]. Our finding that ACC connectivity with regions from both other networks differentiates patients with different treatment outcomes further supports this idea and highlights the potential of this feature for predicting early remission in response to pharmacotherapy.

The connectivity pattern of the posterior DMN seed differentiated all three groups, with early remitters showing the strongest connectivity within the posterior DMN and between the PCC seed and other cingulate areas, while non-remitters exhibited the weakest connectivity between these regions, and late remitters showed intermediate connectivity strengths. The finding that both stronger and weaker connectivity within the DMN was associated with (early) remission might be explained by the observation that the anterior and posterior DMN subnetworks respond differently to pharmacotherapy [38]. Li et al. [38] found that DMN connectivity was increased in patients before treatment in both subnetworks, but only decreased with pharmacotherapy in the posterior DMN. This is consistent with the idea that the stronger posterior DMN connectivity we observed in early and late remitters prior to treatment is one of the MDD-related abnormalities that pharmacotherapy has an effect on. Importantly, the fact that we observed this group difference at baseline suggests that PCC connectivity could be a predictor of treatment success.

With the insula as seed (i.e. focusing on the SN), we identified two connectivity patterns: one distinguishing early and late remitters, and one distinguishing remitters (early and late) from non-remitters. The first connectivity pattern included stronger connectivity in late remitters within the SN (between bilateral insulae) and weaker connectivity between the insula seed and regions from the other two networks (e.g. PCC, dlPFC). Connectivity between limbic SN and frontal CCN regions has often been highlighted as showing treatment-relevant effects [6–8]. In fact, it has been proposed that the right anterior insula acts as a switch between the DMN and CCN, and behaves aberrantly in the context of MDD [54]. While our seed was located in the left anterior insula, our finding of weaker connectivity between the insula seed and the other two networks implies that, in our study as well, the interactions between the SN, DMN and CCN were important for remission. This effect might be specifically related to remission to the aripiprazole add-on many late remitters received, as early remitters, who only received escitalopram for the duration of the study, did not show this effect. Our follow-up analysis comparing remitters to escitalopram alone and remitters to combined escitalopram and aripiprazole revealed some overlap with the early/late remitter contrast, most notably showing weaker connectivity between the insula seed and the anterior DMN (i.e. the mPFC) in both late remitters and remitters to combined escitalopram and aripiprazole. The second contrast further highlighted the importance of SN – DMN connectivity for both early and late remission, with stronger connectivity between the insula seed and the mPFC and the left angular gyrus at baseline being associated with remission.

In addition, this second pattern included multiple cerebellar areas whose connectivity with the insula seed differentiated remitters and non-remitters. Although the cerebellum was not the focus of our study, it showed up in most of our analyses as contributing to the differences between patients and controls, and different patient groups. While the cerebellum has traditionally been associated with motor function, this part of the brain is increasingly recognized as a structure playing an important role in a diverse set of processes and disorders, including the pathology and treatment of MDD [55–57]. Indeed, most of our analyses revealed connections between our seeds and regions in the posterior lobe of the cerebellum (lobule VI-IX), which have more specifically been related to non-motor functions (e.g. cognitive and affective; [55]). The remitter/non-remitter contrast in insula connectivity, which showed the most prominent involvement of the cerebellum in our analyses, revealed greater connectivity between the insula seed and lobule VI, VIIA (crus I & II) and VIII of the cerebellum. Lobule VI has been associated with affective processing, and crus I and II of lobule VIIA have been related to the DMN and CCN, once again pointing towards the importance of networks involved with emotion processing and mood regulation for antidepressant treatment effects.

Interestingly, the analysis with the dlPFC as the seed region (from the CCN) did not reveal any differences among the three patient groups. Previous findings of CCN connectivity have been inconsistent, with some proposing that MDD-related alterations in CCN connectivity might not be sensitive to treatment with antidepressant medication [7]. An alternative interpretation, in line with the fact that CCN regions did show up in our analyses with seeds from the other networks, is that CCN connectivity is only weakly related to pharmacotherapeutic effects (e.g. through its connections with other networks).

To test the stability of our findings, we performed post-hoc analyses for the three seed regions (ACC, PCC and insula) that revealed differences between early-late- and/or non-remitters on the largest single-site dataset to exclude the possibility that our findings might have been unduly influenced by inter-site variability (e.g. due to data being collected using different scanners). Half of the contrasts survived significance testing with correction for multiple comparisons, and all showed trends toward significance and revealed similar patterns of connectivity as observed in the main analyses. These findings suggest that the baseline differences we observed between our three patient groups have a degree of stability, are not dependent on a specific site/scanner setup and are therefore more likely to generalize to other samples. The lack of significance of part of our post-hoc tests is likely related to the smaller sample size used in these analyses.

While our study addressed certain issues relating to reproducibility and generalizability, such as sample size and scanner differences, other factors fell outside the scope of this paper. For example, individual variation in the expression of MDD and response to antidepressant treatment among patients has also been proposed to play a role in the difficulty of finding robust (neuroimaging) biomarkers [6, 58]. Although the bootstrapping procedure in PLS analyses ensures that the identified patterns of connectivity show a degree of stability across participants, this approach is not designed to determine if these findings might apply at an individual patient level. Thus, exploring whether these potential markers of treatment outcome translate to individual patients would be a worthwhile next step towards clinical applications. Similarly, other sources of variance, such as demographic and clinical characteristics and the use of different methodology, have been related to variable findings [5, 6, 58], and could be explored in more detail in future studies.

## Limitations

Our study had several limitations that deserve consideration. First, the treatment regimen differed between early remitters (by definition recipients of escitalopram monotherapy), and among late and non-remitters, who received either escitalopram alone or with adjunctive aripiprazole. While early remitters only received escitalopram, both late and non-remitters included individuals who received either escitalopram alone, or escitalopram and aripiprazole together, making it hard to disentangle specific medication effects. While suboptimal, this is common and difficult to avoid in clinical research on MDD, as the use of multiple medications and individually adjusted regimens are standard treatment practice.

Second, connectivity can be quantified in multiple ways, and using different approaches has been related to the variable findings reported in the literature. We chose seed-based correlation because it is the most commonly used method, and it has shown a greater degree of stability when compared to other metrics of connectivity [59], but it does have its own disadvantages. For example, by correlating a few seeds selected a priori with the rest of the voxels in the brain, our analyses focused on a limited set of connections in the brain, meaning that relevant connections could have been missed. In addition, even within the category of seed-based correlation there are different approaches to calculate connectivity, and the one we used (across-subject cross-correlation) is different from the one employed by Fiecas and colleagues [59] (within-subject cross-correlation, see [60] for details on the difference between these two approaches). Lastly, we selected the ACC as the anterior DMN seed, because this area appears to play a key role in the pathology and treatment of MDD and is most commonly selected as anterior DMN seed in seed-based connectivity studies of MDD [4, 50]. However, this region might only be included in the DMN in patients with MDD [9], and might therefore have skewed our comparison of MDD and control participants for this seed.

## Conclusion

Here we replicated and extended abnormal patterns of fMRI connectivity in patients with MDD compared to controls, as well as connectivity characteristics measured before the start of treatment associated with remission status after 16 weeks of antidepressant treatment. Our replication of connectivity differences between patients and controls confirmed the importance of the DMN, SN and CCN for MDD pathology, and revealed additional stronger connections between the SN and CCN in patients. Our patient group analyses further revealed multiple patterns of connectivity within and between the DMN, SN and CCN that were associated with remission status. Interestingly, different seeds highlighted different group characteristics, indicating that multiple networks need to be considered to get a full picture of the connectivity differences between early, late and non-remitters to pharmacotherapy. Specifically, ACC connectivity revealed a pattern that was specific to early remitters, while insula connectivity revealed a late-remitter and remitter effect and PCC connectivity distinguished all three groups. Although reproducibility and generalizability are still major concerns in current neuroimaging research, the fact that we were able to replicate previous single-site findings in a larger, multi-site sample (with MRI data collected from different Tesla scanners), and demonstrate stability in the patterns differentiating patients based on remission status in a single-site subsample of our data is encouraging and supports the potential of our findings for future clinical use.

## Data Availability

The data analyzed in this paper are not publicly available, but may be made available if permission is granted by the CAN-BIND Investigator's Team and Ontario Brain Institute upon reasonable request.

## Acknowledgements

We would like to thank Keith Ho for his help with organizing the clinical data, and all the patients, controls, and health care workers involved in this project for the generous contribution of their time and their participation in this work.

## Author contributions

GW: Data analysis and interpretation, writing (original draft), writing (review, editing, and approval); JKH: Data collection, data curation, writing (review, editing, and approval); SH: Data collection, data curation, writing (review, editing, and approval); ADD: Data collection, data curation, writing (review, editing, and approval); MZ: Data collection, data curation, writing (review, editing, and approval); SRA: Data curation, writing (review, editing, and approval); RM: Study concept, data collection, writing(review, editing, and approval); RWL: Study concept, data collection, writing (review, editing, and approval); BNF: Study concept, data collection, data curation, writing (review, editing, and approval); GBH: Study concept, data collection, data curation, writing (review, editing, and approval); DJM: Study concept, data collection, data curation, writing (review, editing, and approval); SR: Study concept, data collection, data curation, writing (review, editing, and approval); SHK: Study concept, data collection, writing (review, editing, and approval); SCS: Data curation, writing (review, editing, and approval); GMM: Study concept, data collection, data curation; ABP: Data analysis, writing (original draft), writing (review, editing, and approval).

