## Supplementary material for "Baseline functional connectivity in resting state networks associated with depression and remission status after 16 weeks of pharmacotherapy: A CAN-BIND Report": Table S1

**Table S1**. Regions Showing Distinct Connectivity with the Anterior Cingulate Cortex in Patients with MDD and Controls Automatically Labeled Using the AAL Atlas

| **Contrast** | **Cluster** | **Cluster size** | **Area** | **Number of voxels** | **Peak BSR** | **Peak MNI coordinates**  **x y z** | | |
| --- | --- | --- | --- | --- | --- | --- | --- | --- |
| LV1:  P > C | 1 | 123 | L medial superior frontal gyrus | 16 | 6.71 | 0 | 56 | 0 |
|  |  |  | R medial superior frontal gyrus | 22 | 7.53 | 4 | 52 | 4 |
|  |  |  | L medial superior frontal gyrus, orbital part | 22 | 13.37 | 0 | 56 | -8 |
|  |  |  | R medial superior frontal gyrus, orbital part | 31 | 9.37 | 4 | 48 | -4 |
|  |  |  | L gyrus rectus | 9 | 6.56 | -8 | 52 | -16 |
|  |  |  | L anterior cingulate/paracingulate gyri | 18 | 6.09 | 0 | 48 | -4 |
|  | 2 | 13 | R dorsolateral superior frontal gyrus | 13 | 4.51 | 20 | 44 | 44 |
|  | 3 | 24 | L medial superior frontal gyrus | 13 | 4.35 | -4 | 52 | 24 |
|  |  |  | R medial superior frontal gyrus | 10 | 3.23 | 4 | 56 | 28 |
|  | 4 | 16 | L middle frontal gyrus, orbital part | 8 | 4.03 | -28 | 48 | -12 |
|  |  |  | L inferior frontal gyrus, orbital part | 8 | 3.87 | -32 | 48 | -16 |
|  | 5 | 21 | R angular gyrus | 19 | 3.95 | 44 | -68 | 40 |
|  | 6 | 14 | L temporal pole: superior temporal gyrus | 11 | 3.93 | -40 | 20 | -24 |
|  | 7 | 20 | L cuneus | 4 | 3.32 | -12 | -60 | 24 |
|  |  |  | L precuneus | 15 | 3.67 | -8 | -56 | 24 |
|  | 8 | 31 | L angular gyrus | 21 | 3.49 | -52 | -64 | 32 |
|  |  |  | L middle temporal gyrus | 9 | 3.21 | -52 | -60 | 20 |
|  | 9 | 21 | L middle cingulate/paracingulate gyri | 13 | 2.91 | 0 | -36 | 40 |
|  |  |  | R middle cingulate/paracingulate gyri | 3 | 3.24 | 4 | -40 | 40 |
|  |  |  | R precuneus | 3 | 2.65 | 4 | -44 | 40 |
|  | 10 | 11 | R rolandic operculum | 3 | 2.65 | 40 | -20 | 16 |
|  |  |  | R insula | 7 | 2.89 | 40 | -16 | 4 |
| LV1:  P < C | 11 | 23 | L calcarine fissure | 10 | -4.01 | -4 | -84 | -4 |
|  |  |  | L lingual gyrus | 4 | -3.14 | -12 | -88 | -4 |
|  |  |  | R lingual gyrus | 6 | -3.50 | 4 | -72 | -4 |
|  | 12 | 13 | R supramarginal gyrus | 12 | -3.86 | 52 | -40 | 36 |
|  | 13 | 23 | R superior parietal gyrus | 12 | -3.01 | 28 | -52 | 56 |
|  |  |  | R inferior parietal gyrus | 11 | -3.37 | 36 | -48 | 52 |
|  | 14 | 17 | L middle frontal gyrus | 13 | -3.10 | -40 | 36 | 28 |
|  |  |  | L inferior frontal gyrus, triangular part | 3 | -2.93 | -36 | 36 | 24 |

*Note.* Only the regions included in the 10-15 most stable clusters per LV are presented in this table (|Bootstrap Ratios| ≥ 2.1, Cluster Size ≥ 10). P = patients, C = controls, BSR = bootstrap ratio, R = right, L = left.

**Table S2**. Regions Showing Distinct Connectivity with the Posterior Cingulate Cortex in Patients with MDD and Controls Automatically Labeled Using the AAL Atlas

| **Contrast** | **Cluster** | **Cluster size** | **Area** | **Number of voxels** | **Peak BSR** | **Peak MNI coordinates**  **x y z** | | |
| --- | --- | --- | --- | --- | --- | --- | --- | --- |
| LV1:  P > C | 1 | 45 | R postcentral gyrus | 13 | 3.43 | 44 | -36 | 60 |
|  |  |  | R superior parietal gyrus | 13 | 4.77 | 36 | -48 | 56 |
|  |  |  | R inferior parietal gyrus | 18 | 3.50 | 48 | -40 | 56 |
|  | 2 | 15 | L supplementary motor area | 7 | 3.03 | 0 | 8 | 48 |
|  |  |  | R supplementary motor area | 8 | 4.05 | 4 | 8 | 52 |
|  | 3 | 25 | L medial superior frontal gyrus | 8 | 3.07 | 0 | 36 | 36 |
|  |  |  | R anterior cingulate/paracingulate gyri | 6 | 2.68 | 4 | 40 | 28 |
|  |  |  | R middle cingulate/paracingulate gyri | 9 | 3.87 | 4 | 28 | 36 |
|  | 4 | 15 | R dorsolateral superior frontal gyrus | 12 | 3.66 | 24 | 32 | 52 |
|  |  |  | R middle frontal gyrus | 3 | 2.80 | 24 | 32 | 44 |
|  | 5 | 18 | R middle frontal gyrus | 16 | 3.43 | 40 | 12 | 40 |
|  | 6 | 10 | L precentral gyrus | 9 | 3.10 | -60 | 12 | 32 |
|  | 7 | 13 | L inferior frontal gyrus, opercular part | 8 | 3.08 | -60 | 16 | 4 |
|  |  |  | L inferior frontal gyrus, triangular part | 3 | 2.29 | -60 | 20 | 16 |
|  | 8 | 15 | R precentral gyrus | 13 | 3.01 | 48 | 0 | 36 |
|  | 9 | 14 | L middle frontal gyrus | 13 | 2.87 | -28 | 8 | 48 |
| LV1:  P < C | 10 | 11 | L precuneus gyrus | 4 | -2.61 | -16 | -60 | 36 |
|  | 11 | 10 | R middle temporal gyrus | 4 | -2.89 | 52 | -12 | -24 |
|  |  |  | R inferior temporal gyrus | 6 | -3.62 | 56 | -8 | -28 |
|  | 12 | 10 | R lobule VIII of cerebellar hemisphere | 8 | -3.51 | 20 | -56 | -48 |

*Note.* Only the regions included in the 10-15 most stable clusters per LV are presented in this table (|Bootstrap Ratios| ≥ 2.0, Cluster Size ≥ 10). P = patients, C = controls, BSR = bootstrap ratio, R = right, L = left.

**Table S3**. Regions Showing Distinct Connectivity with the Insula in Patients with MDD and Controls Automatically Labeled Using the AAL Atlas

| **Contrast** | **Cluster** | **Cluster size** | **Area** | **Number of voxels** | **Peak BSR** | **Peak MNI coordinates**  **x y z** | | |
| --- | --- | --- | --- | --- | --- | --- | --- | --- |
| LV1:  P > C | 1 | 18 | L precentral gyrus | 6 | 3.45 | -60 | 0 | 36 |
|  |  |  | L postcentral gyrus | 12 | 3.48 | -52 | -8 | 44 |
|  | 2 | 26 | L inferior frontal gyrus, opercular part | 6 | 3.22 | -60 | 16 | 20 |
|  |  |  | L inferior frontal gyrus, triangular part | 20 | 3.07 | -52 | 24 | 20 |
|  | 3 | 11 | L middle frontal gyrus | 11 | 2.76 | -44 | 20 | 44 |
| LV1:  P < C | 4 | 12 | L fusiform gyrus | 7 | -2.66 | -32 | -56 | -20 |
|  |  |  | L lobule VI of cerebellar hemisphere | 5 | -4.05 | -32 | -60 | -20 |
|  | 5 | 19 | L Lobule IX of cerebellar hemisphere | 6 | -4.00 | -4 | -60 | -44 |
|  |  |  | Lobule IX of vermis | 7 | -3.07 | -4 | -56 | -32 |
|  | 6 | 17 | L insula | 13 | -3.61 | -36 | 20 | 4 |
|  | 7 | 10 | R fusiform gyrus | 4 | -3.35 | 36 | -68 | -20 |
|  |  |  | R lobule VI of cerebellar hemisphere | 3 | -2.34 | 32 | -68 | -20 |
|  | 8 | 16 | R calcarine fissure | 6 | -3.20 | 8 | -60 | 12 |
|  |  |  | R cuneus | 3 | -2.84 | 8 | -60 | 20 |
|  |  |  | R precuneus | 5 | -2.80 | 4 | -60 | 20 |
|  | 9 | 15 | L calcarine fissure | 12 | -3.12 | -8 | -92 | -8 |
|  | 10 | 10 | L calcarine fissure | 4 | -2.54 | -24 | -68 | 12 |
|  | 11 | 10 | L lingual gyrus | 8 | -2.70 | 12 | -88 | -12 |
|  | 12 | 14 | L lingual gyrus | 7 | -3.04 | -20 | -48 | -8 |
|  |  |  | L fusiform gyrus | 6 | -2.85 | -16 | -40 | -16 |
|  | 13 | 12 | R Lobule VIII of cerebellar hemisphere | 5 | -2.92 | 20 | -56 | -44 |
|  |  |  | R Lobule IX of cerebellar hemisphere | 3 | -2.81 | 20 | -48 | -44 |
|  | 14 | 13 | R lingual gyrus | 9 | -2.87 | 20 | -48 | -8 |

*Note.* Only the regions included in the 10-15 most stable clusters per LV are presented in this table (|Bootstrap Ratios| ≥ 2.0, Cluster Size ≥ 10). P = patients, C = controls, BSR = bootstrap ratio, R = right, L = left.

**Table S4**. Regions Showing Distinct Connectivity with the Dorsolateral Prefrontal Cortex in Patients with MDD and Controls Automatically Labeled Using the AAL Atlas

| **Contrast** | **Cluster** | **Cluster size** | **Area** | **Number of voxels** | **Peak BSR** | **Peak MNI coordinates**  **x y z** | | |
| --- | --- | --- | --- | --- | --- | --- | --- | --- |
| LV1:  P > C | 1 | 12 | R middle frontal gyrus | 12 | 4.05 | 44 | 12 | 40 |
|  | 2 | 11 | R inferior parietal gyrus | 9 | 3.93 | 36 | -52 | 52 |
|  | 3 | 17 | L lingual gyrus | 9 | 3.47 | -8 | -80 | -4 |
|  |  |  | Lobule VI of vermis | 3 | 2.78 | 0 | -72 | -8 |
|  | 4 | 12 | L fusiform gyrus | 7 | 3.02 | -28 | -36 | -24 |
|  |  |  | L lobule IV, V of cerebellar hemisphere | 3 | 2.89 | -28 | -40 | -24 |
| LV1:  P < C | 5 | 13 | L medial superior frontal gyrus | 8 | -4.37 | -8 | 28 | 40 |
|  | 6 | 16 | R lobule VIII of cerebellar hemisphere | 8 | -4.34 | 28 | -52 | -44 |
|  | 7 | 29 | L precentral gyrus | 3 | -2.61 | -44 | 8 | 32 |
|  |  |  | L middle frontal gyrus | 15 | -3.62 | -44 | 40 | 28 |
|  |  |  | L inferior frontal gyrus, opercular part | 6 | -3.04 | -44 | 20 | 32 |
|  |  |  | L inferior frontal gyrus, triangular part | 5 | -2.77 | -44 | 20 | 28 |
|  | 8 | 11 | R crus I of cerebellar hemisphere | 5 | -2.62 | 16 | -72 | -32 |
|  |  |  | R crus II of cerebellar hemisphere | 5 | -3.51 | 24 | -72 | -40 |
|  | 9 | 33 | L inferior frontal gyrus, opercular part | 3 | -2.74 | -44 | 12 | 8 |
|  |  |  | L inferior frontal gyrus, triangular part | 12 | -3.40 | -36 | 20 | 8 |
|  |  |  | L insula | 16 | -3.30 | -32 | 24 | 8 |
|  | 10 | 10 | L superior frontal gyrus | 6 | -2.76 | -24 | 60 | 8 |
|  |  |  | L middle frontal gyrus | 3 | -3.05 | -24 | 52 | 8 |
|  | 11 | 15 | R insula | 3 | -3.00 | 40 | 4 | 12 |
|  |  |  | R putamen | 8 | -2.96 | 28 | 8 | 8 |

*Note.* Only the regions included in the 10-15 most stable clusters per LV are presented in this table (|Bootstrap Ratios| ≥ 2.0, Cluster Size ≥ 10). P = patients, C = controls, BSR = bootstrap ratio, R = right, L = left.

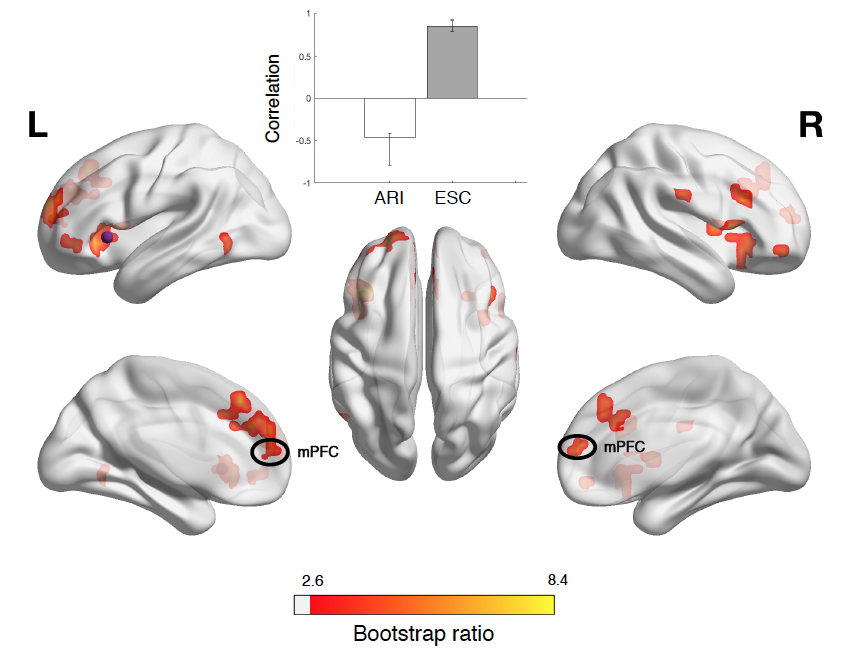

**Figure S1**. Results from PLS analysis comparing remitters to escitalopram alone and remitters to combined escitalopram and aripiprazole in terms of connectivity between the insula ROI (purple sphere) and all other voxels, projected onto a smoothed cortical surface. The correlation bar graph shows group-dependent differences in the correlation with the seed voxels of the areas identified in the brain image. The error bars indicate the 95% confidence intervals derived from bootstrap estimation. The brain image illustrates the areas that expressed this contrast most stably across participants, as determined by bootstrapping. Only the 10-15 clusters (>10 voxels) with the highest bootstrap ratios (|BSR| ≥ 2.6) are presented, and the cerebellum is not included. Red/yellow clusters indicate stronger connectivity in remitters to escitalopram alone, compared to remitters to combined escitalopram and aripiprazole. ARI = remitters to combined escitalopram and aripiprazole, SC = remitters to escitalopram alone, mPFC = medial prefrontal cortex.

**Table S5.** Regions Showing Distinct Connectivity with the Insula ROI in Patients Who Remitted to Escitalopram Alone compared to Patients Who Remitted to Combined Escitalopram and Aripiprazole Automatically Labeled Using the AAL Atlas

| **Contrast** | **Cluster** | **Cluster size** | **Area** | **Number of voxels** | **Peak BSR** | **Peak MNI coordinates**  **x y z** | | |
| --- | --- | --- | --- | --- | --- | --- | --- | --- |
| LV1: ESC > ESC+ARI | 1 | 81 | L inferior frontal gyrus, opercular part | 5 | 4.14 | -40 | 8 | 8 |
|  |  |  | L inferior frontal gyrus, triangular part | 33 | 6.81 | -40 | 20 | 8 |
|  |  |  | L inferior frontal gyrus, orbital part | 16 | 6.33 | -36 | 28 | -4 |
|  |  |  | L insula | 23 | 8.42 | -40 | 20 | 4 |
|  | 2 | 49 | L medial superior frontal gyrus | 19 | 6.15 | -4 | 32 | 44 |
|  |  |  | R medial superior frontal gyrus | 6 | 4.93 | 4 | 32 | 44 |
|  |  |  | L anterior cingulate/paracingulate gyri | 4 | 3.55 | -12 | 28 | 28 |
|  |  |  | R anterior cingulate/paracingulate gyri | 4 | 4.39 | 4 | 28 | 28 |
|  |  |  | L middle cingulate/paracingulate gyri | 4 | 3.69 | -8 | 20 | 36 |
|  |  |  | R middle cingulate/paracingulate gyri | 9 | 4.06 | 4 | 32 | 36 |
|  | 3 | 48 | L medial superior frontal gyrus | 31 | 5.45 | -12 | 52 | 12 |
|  |  |  | R medial superior frontal gyrus | 5 | 4.12 | 4 | 52 | 16 |
|  |  |  | L anterior cingulate/paracingulate gyri | 6 | 4.22 | -4 | 40 | 24 |
|  |  |  | R anterior cingulate/paracingulate gyri | 3 | 3.89 | 8 | 48 | 16 |
|  | 4 | 37 | L superior frontal gyrus | 21 | 5.40 | -16 | 60 | 32 |
|  |  |  | L middle frontal gyrus | 15 | 3.90 | -36 | 44 | 16 |
|  | 5 | 51 | R inferior frontal gyrus, opercular part | 11 | 5.16 | 52 | 16 | 8 |
|  |  |  | R inferior frontal gyrus, triangular part | 7 | 4.04 | 52 | 20 | 4 |
|  |  |  | R inferior frontal gyrus, orbital part | 7 | 4.14 | 36 | 24 | -8 |
|  |  |  | R insula | 22 | 5.12 | 28 | 20 | -20 |
|  | 6 | 12 | R middle frontal gyrus, orbital part | 5 | 3.65 | 40 | 48 | -8 |
|  |  |  | R inferior frontal gyrus, orbital part | 6 | 4.85 | 44 | 48 | -12 |
|  | 7 | 15 | R insula | 9 | 4.63 | 36 | 4 | 8 |
|  | 8 | 20 | L middle frontal gyrus | 3 | 4.08 | -48 | 48 | 0 |
|  |  |  | L middle frontal gyrus, orbital part | 4 | 4.37 | -48 | 48 | -4 |
|  |  |  | L inferior frontal gyrus, orbital part | 9 | 4.27 | -52 | 32 | -8 |
|  | 9 | 12 | R inferior frontal gyrus, opercular part | 5 | 3.94 | 44 | 16 | 28 |
|  |  |  | R inferior frontal gyrus, triangular part | 7 | 4.27 | 44 | 20 | 24 |
|  | 10 | 11 | R supramarginal gyrus | 9 | 3.96 | 60 | -20 | 28 |
|  | 11 | 15 | L putamen | 13 | 3.92 | -28 | 0 | -4 |
|  | 12 | 14 | L middle temporal gyrus | 11 | 3.88 | -52 | -56 | 0 |
|  |  |  | L inferior temporal gyrus | 3 | 3.58 | -52 | -60 | -8 |

*Note.* Only the regions included in the 10-15 most stable clusters per LV are presented in this table (|Bootstrap Ratios| ≥ 2.6, Cluster Size ≥ 10). ESC = patients who remitted to escitalopram alone, ARI = patients who remitted to combined escitalopram and aripiprazole, BSR = bootstrap ratio, R = right, L = left.
